# Age trajectories of infant mortality among Palestinian refugees born in 2010–2020: evidence from electronic health records

**DOI:** 10.64898/2026.09.22.26363685

**Authors:** Selin Köksal, Yu Chen, Zeina Jamaluddine, Seita Akihiro, Francesco Checchi, Oona M. R. Campbell, José Manuel Aburto

**Author notes:** These authors contributed equally (Selin Köksal, Yu Chen).

## Abstract

**Objectives:** To estimate age trajectories of infant mortality among Palestinian refugees using electronic health records, addressing the scarcity of reliable mortality data in forced displacement settings.

**Setting:** Palestinian refugees living in Gaza, West Bank, Jordan, Syria and Lebanon.

**Design:** We fitted a Bayesian Negative Binomial model to observed death counts, using a flexible parametric function (a shifting power function) to capture the shape of the infant mortality curve, across strata to infer age trajectories of infant mortality.

**Participants:** Palestinian refugees born in 2010–2020 and registered with UNRWA health services in Gaza, West Bank, Jordan, Syria and Lebanon. The sample was stratified by sex, field, gestational age and birth cohort.

**Main outcome measures:** Survival rate to age 1.

**Results:** Overall, the survival rate to age 1 was 98.64%, with slightly lower rates for males (98.57%) than females (98.68%). Across settings, West Bank refugees had the lowest infant mortality (99.08% survival rate), and Jordan refugees had the highest (98.22% survival rate). Results by gestational age showed a 98.85% survival rate at age 1 among term births compared to a 73.32% survival rate among very preterm births. Survival rates by cohort (births up to 2015 or after) were 98.78% and 98.42%, respectively.

**Conclusion:** Our findings highlight the potential of electronic health records for mortality monitoring in forced displacement settings as they reveal consistent mortality trajectories by sex and gestational age, while identifying meaningful variation across refugee settings and subgroups.

**Summary boxes:** *What is already known in this topic:* We searched PubMed and Google Scholar for articles published between Jan 1, 2000, and Feb 20, 2026, in English, using the terms “Palestinian refugees,” “infant mortality,” “neonatal mortality,” and “UNRWA.” We found a handful of studies estimating infant mortality among Palestinian refugees, mostly based on household surveys (eg, MICS, national statistical office surveys) or UNRWA surveys using preceding birth techniques. The preceding birth technique has notable limitations, including recall bias, small sample sizes, and selection bias. The majority of these studies focus exclusively on Palestinian refugees residing in Gaza. Estimates for Gaza suggest a sustained decline, from around 127 deaths per 1,000 births in 1960 to 20.2 per 1,000 births in 2006, with a slowdown thereafter and an increase in more recent years. A combined estimate (excluding Syria) across four settings where Palestinian refugees reside measures infant mortality at 18 per 1,000 births in 2011.

*What this study adds:* This study uses a unique data linkage bringing together UNRWA’s obstetric and child health records, providing more extensive information and a larger sample size than previous studies. This is also the first study to assess the data quality of these electronic records and to estimate age trajectories of infant mortality among Palestinian refugees. We developed a statistical strategy that estimated infant mortality trajectories covering all five UNRWA fields (Gaza, Jordan, Lebanon, Syria, and the West Bank) for children born in 2010-2020, and examined differences across subpopulations by sex, field, gestational age and cohort.

## Introduction

Infant mortality is a key global health indicator. Yet, data on refugees and other marginalized groups are often sparse and underrepresented, making it difficult to estimate mortality reliably. Retrospective household surveys relying on the SMART (Standardised Monitoring and Assessment of Relief and Transitions) methodology as well as DHS and MICS surveys remain an important source of information on infant mortality for refugee populations. However, they can be slow and costly to produce and often contain limited sample sizes for more granular analysis [1]. Furthermore, during and after conflict periods, survey fieldwork is frequently disrupted, undermining the capacity to monitor populations and inform humanitarian response [2]. The recent expansion of electronic health records in epidemiological research offers a promising alternative for tracking refugee health both during and outside of conflict [3], providing large-scale and cost-effective information that are critical in resource-scarce settings [4].

In this study, we focus on the world’s largest protracted refugee population: Palestinian refugees, who number almost six million and represent around one fifth of the global refugee population [5]. Palestinian refugees have experienced displacement and marginalisation since 1948, and today most of them reside across five settings: Jordan, Lebanon, Syria, the West Bank, and Gaza. Reliable estimates of basic mortality indicators for this population remain scarce, limiting the ability to monitor health and mortality trajectories. To contribute to a better understanding of Palestinian refugee health, we draw on a unique dataset of linked electronic medical records covering nearly one million Palestinian refugees born between 2010 and 2020 [6], and provide the first comprehensive analysis of data quality and age-specific infant mortality trajectories in this population

The limited prior evidence on infant mortality among Palestinian refugees draws on household surveys, such as MICS, those conducted by national statistical offices, and on surveys administered by the United Nations Relief and Works Agency for Palestine Refugees in the Near East (UNRWA), the main agency providing health and social care for refugees. These sources have produced infant mortality estimates periodically, though not systematically. Estimates indicate a sustained decline in Gaza [7] from around 127 deaths per 1,000 births in 1960 to 20.2 per 1,000 births in 2006 [8], albeit with a slowdown in progress since then [9] and increased infant mortality in more recent years [8, 10]. A similar pattern of decline followed by stalling was also observed across other settings where Palestinian refugees reside (see Supplementary Table 1 for a summary of previous estimates), with an aggregate (excluding Syria) infant mortality rate of 18 per 1,000 births in 2011 [11]. These studies mostly rely on the preceding birth technique, in which mothers with at least two children are interviewed about the survival of the child born just before their most recent birth. While this method has several strengths in data scarce contexts, it carries limitations such as recall bias, limited sample size, and selection bias [12] that can be mitigated by using health records.

## Methods

### Data

We used previously collected, and anonymised data from UNRWA’s medical records covering nearly one million Palestinian refugee children born between 1 January 2010 and 31 December 2020, and linked these with their mother’s obstetric records. Palestinian refugees are defined as all descendants of Palestinian refugee males, whose normal place of residence was Palestine between June 1st 1946 and May 15th 1948, and who lost both their homes and means of livelihood as a result of the 1948 conflict [6, 13]. UNRWA provides free primary healthcare services to Palestinian refugees in Gaza, Jordan, Lebanon, Syria, and West Bank, including antenatal care services and maintains an electronic administrative database recording all visits to UNRWA clinics. UNRWA does not directly provide childbirth services (except one hospital in West Bank) [14] but collects obstetric records. The linkage across obstetric, health, and education records was made possible by a unique ID assigned to each registered refugee [6]. For a given setting, UNRWA data captures only refugees who use UNRWA services for healthcare. This does not imply that all registered refugees live in camps nor that camp residency determines service use. Palestinian refugees living outside camps can still access UNRWA services. Regardless of place of residence, Palestinian refugees tend to use UNRWA services on grounds of eligibility and cost. However, the coverage of obstetric records is not exhaustive and antenatal care (ANC) utilisation varies substantially by field. The proportion of pregnant Palestinian refugees attending ANC at UNRWA services are estimated at 73% in Gaza, 49% in West Bank, 35% in Jordan in 34% in Syria in 2019, and no estimate is available for Lebanon [14]. UNRWA’s electronic medical records capture pregnancy outcome during postnatal care, so we expect the population captured in the obstetric records to closely align with ANC attendees. In Supplementary Figure 1, we present a flowchart summarising the population covered in UNRWA data.

### Data quality

Infant deaths, particularly neonatal deaths, often do not appear on healthcare records because newborns who die very early are not brought to child healthcare services. As a result, the percentage of infant deaths not recorded can be as high as 80% [6]. A total of 14,009 infant and child deaths were counted in mother obstetric records and child health records among 972,743 births (see Supplementary Table 2 for a breakdown by sex, field, gestational age, and cohort).

The age at death was recorded in unstandardised free-text fields within the obstetric records by nurses, requiring extensive data cleaning to identify these deaths and establish their timing. Uncertainty in dates was partially resolved by setting the latest recorded health service access as the earliest-possible date, and by treating the ambiguous time unit abbreviation ‘m’ as months if the child had a history of health service visits consistent with this time frame, or minutes otherwise. 16 cases had a recorded death date that preceded the birth date. We treated these as misrecordings, assuming the dates had been swapped (dates are variously recorded as MMDDYYYY or DDMMYY depending on the registering provider). For a substantial share of death records we were left with either a date interval (n = 6,122, 44%) or no clear age at death information (n = 2,981, 21%); for the remaining 4,906 (35%), age at death was recorded unambiguously.

To assess data quality prior to imputation, we conducted an age heaping analysis for infant deaths with complete age-at-death information (see Data quality section in Supplementary Material). Evidence for age heaping informed our modelling strategy, which accounts for heaping and assigns greater weight to deaths in the earliest days after birth, as described in the Statistical analysis section.

### Imputation

To resolve the age-at-death uncertainty for the 6,122 interval-censored and 2,981 fully unknown cases, we fit a piecewise exponential survival model to the subset of the data comprising surviving children and deceased children with unambiguously recorded mortality dates and use it to predict the age at death of those cases with interval censoring. This approach preserved the observed age pattern of mortality and avoided cruder forms of imputation (e.g., midpoint imputation). We also imputed 1609, (11.5%) missing sex values by assuming a birth sex ratio of 1.05 females to males.

### Statistical analysis

We developed a Bayesian regression model to estimate daily infant mortality rates during the first year of life, assuming a negative binomial distribution to account for overdispersion (parameter *φ*) in the observed death counts. In the Bayesian framework, we incorporated demographic theory to model mortality risk shape as a parametric power-law based on previous evidence [15]. With this strategy, our model addressed age-heaping and noisy data issues. We denote the infant death counts by 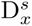, where *x* is the number of days since birth. The superscript *s* represents stratification by sex, field setting, gestational age groups, or cohort group. The exposure by day is denoted by 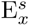.

We modelled the expected death counts, 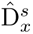, using a log-link function (Equation (1b)) to ensure non-negative values, and used the exposure (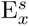) as an offset. To accommodate the highly concentrated mortality counts in the first week since birth, we applied observation-specific weights on the likelihood using a calibration approach (Equations (1c–1e)). We validated our model choice and provided additional details in the Supplementary Materials.

The model is specified as

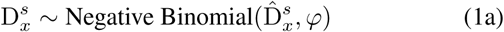

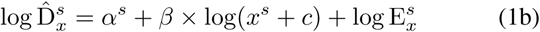

with weighted likelihood

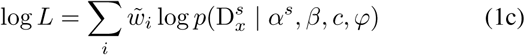

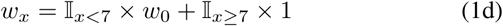

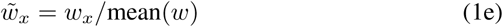

In Equation (1), we assume similar mortality shape across strata *s* with shared slope *β* and location-shifting parameter *c*, consistent with previous evidence [16, 17]. To account for the strata-specific mortality rates, we used fixed effects on the baseline parameter *α^s^*. We compared this assumption with random effects in the Supplementary Materials. Prior distributions were selected with weakly informative distributions based on the default settings [18]. The corresponding prior specifications and sensitivity checks are provided in the Supplementary Materials.

Analyses were conducted for the full sample and stratified by sex, by setting, by gestational age group ([28, 32) weeks as very preterm, [32, 37) weeks as preterm, and [37, 42) weeks as term), and by cohort (births up to 31 December 2015, and after 31 December 2015). In each scenario, we implemented the model with the Hamiltonian Monte Carlo algorithm in `Stan` through `brm` [19] package, using four parallel chains, with a warm-up period of 1,000 iterations and 4,000 sampling iterations. The detailed package versions are provided in the Supplementary Material. Convergence and mixing were confirmed by a maximum Gelman-Rubin diagnostic (*R̂*) and effective sample size. Model adequacy was evaluated using prior predictive checks and posterior predictive simulations.

We compared our model with GLM models used in the literature, highlighting the scalability and robustness of our central model. In addition, we evaluated our choice of weights on the likelihood and the parameter assumption. Further details are provided in the Supplementary Materials.

### Main outputs

From the posterior distribution, we derived three functions: the age-specific hazard, the age-specific survival function, and the cumulative distribution of infant deaths using standard procedures [16].

In Figs 1–4, we present the posterior median and 95% credibility intervals (Cr.I.) of age-specific mortality rates, together with estimates of the other two quantities derived from posterior median mortality rates.

**Fig. 1:**
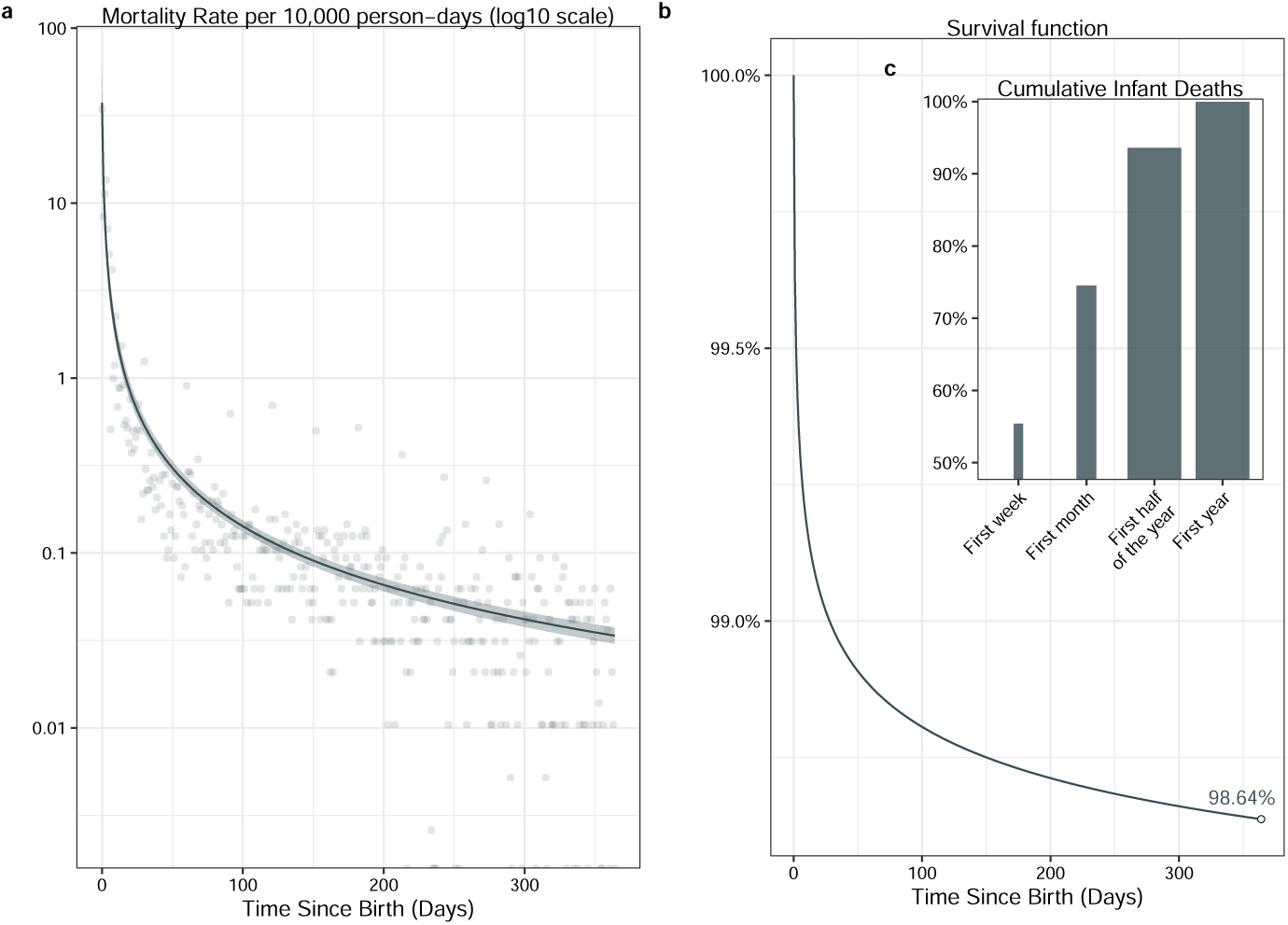
The infant mortality and survival function trends in the first year since birth. **(a).** The estimated infant mortality rate per 10,000 person days with the base-10 log transformation in the first year since birth (x-axis) with the fitted model log D̂*_x_* = −6.00 − 1.12 *×* log(*x* + 0.69) + log(E*_x_*). The posterior median values of mortality rates (−6.00 − 1.12 *×* log(*x* + 0.69)) are shown in line with 95% Credible Interval (Cr.I.) in ribbons. The corresponding Cr.I. are shown in Table 3. The empirical mortality rates are shown in dots. **(b).** The estimated survival function based on the posterior median mortality rates. The probability of survival on day 364 is shown in the circle with the value in text. **(c).** The cumulative probability distributions in terms of infant deaths (bars) in the specific time periods (the first week ([0, 6] days), the first month ([0, 28] days), the first half year ([0, 182] days) and the first year ([0, 364] days)). The widths of the bars indicate the relative lengths of the time periods. The detailed information is shown in Table 1.

**Fig. 2:**
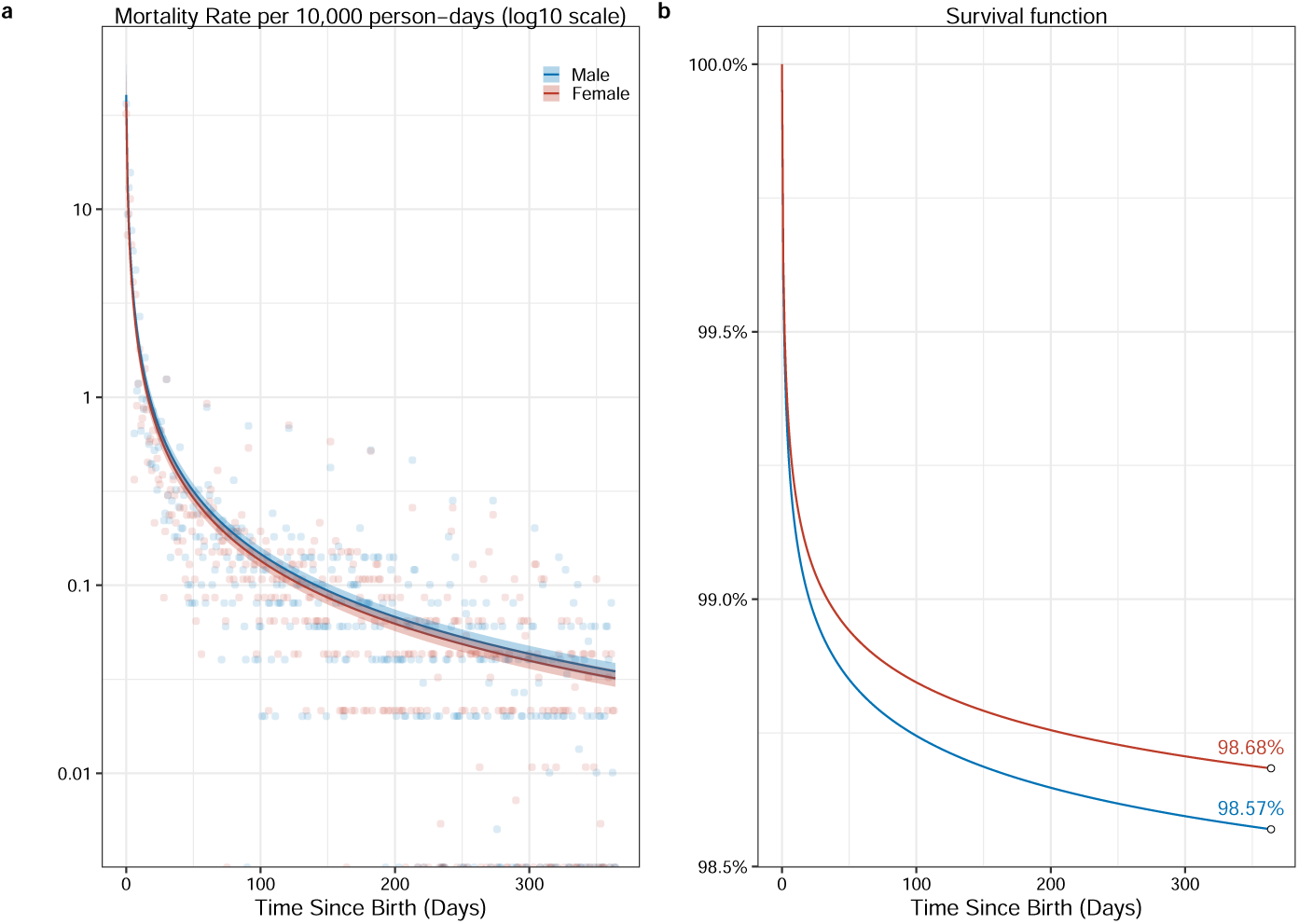
The infant mortality and survival function trends in the first year by sex since birth. **(a).** The estimated infant mortality rate per 10,000 person days with the base-10 log transformation in the first year since birth (x-axis) with the fitted model log D̂*_x_* = *α −* 1.12 *×* log(*x* + 0.66) + log(E*_x_*), with the fix effects *α* for male of −5.97 and female of −6.06. The corresponding Cr.I. are shown in Table 3. The posterior median values of mortality rates are shown in line with 95% Cr.I. in ribbons. The empirical mortality rates are shown in dots. **(b).** showed the same estimates based on the posterior median value of mortality rates by sex (colour).

**Fig. 3:**
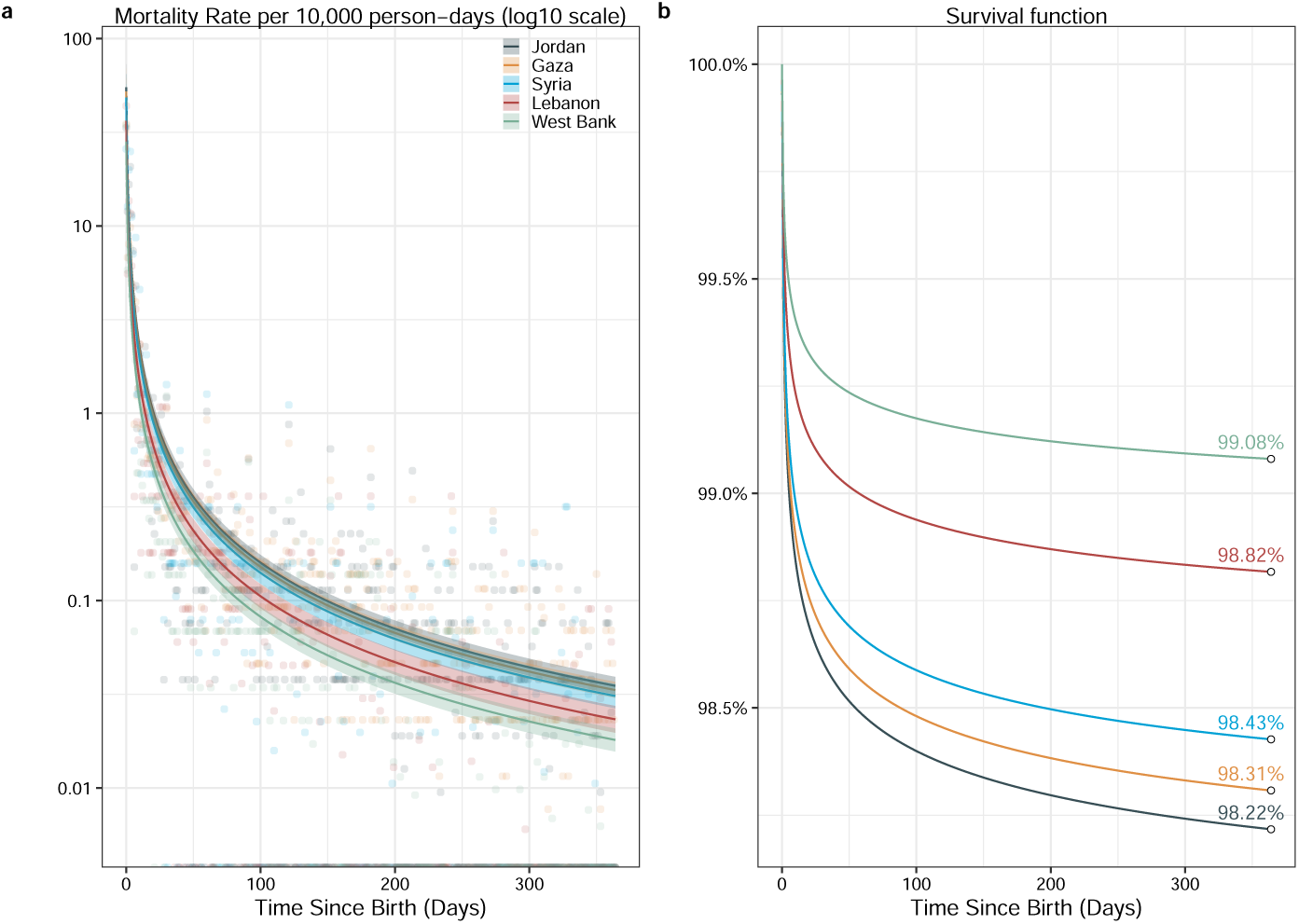
The infant mortality and survival function trends in the first year by sex since birth. **(a).** The estimated infant mortality rate per 10,000 person days with the base-10 log transformation in the first year since birth (x-axis) with the fitted model log D̂*_x_* = *α −* 1.18 *×* log(*x* +0.70) +log(E*_x_*), with the fix effects *α* for Jordan, Gaza, Syria, Lebanon, West Bank of −5.62, −5.68, −5.75, −6.04, −6.29, respectively. The corresponding Cr.I. are shown in Table 3. The posterior median values of mortality rates are shown in line with 95% Cr.I. in ribbons. The empirical mortality rates are shown in dots. **(b).** showed the same estimates based on the posterior median value of mortality rates by setting (colour).

**Fig. 4:**
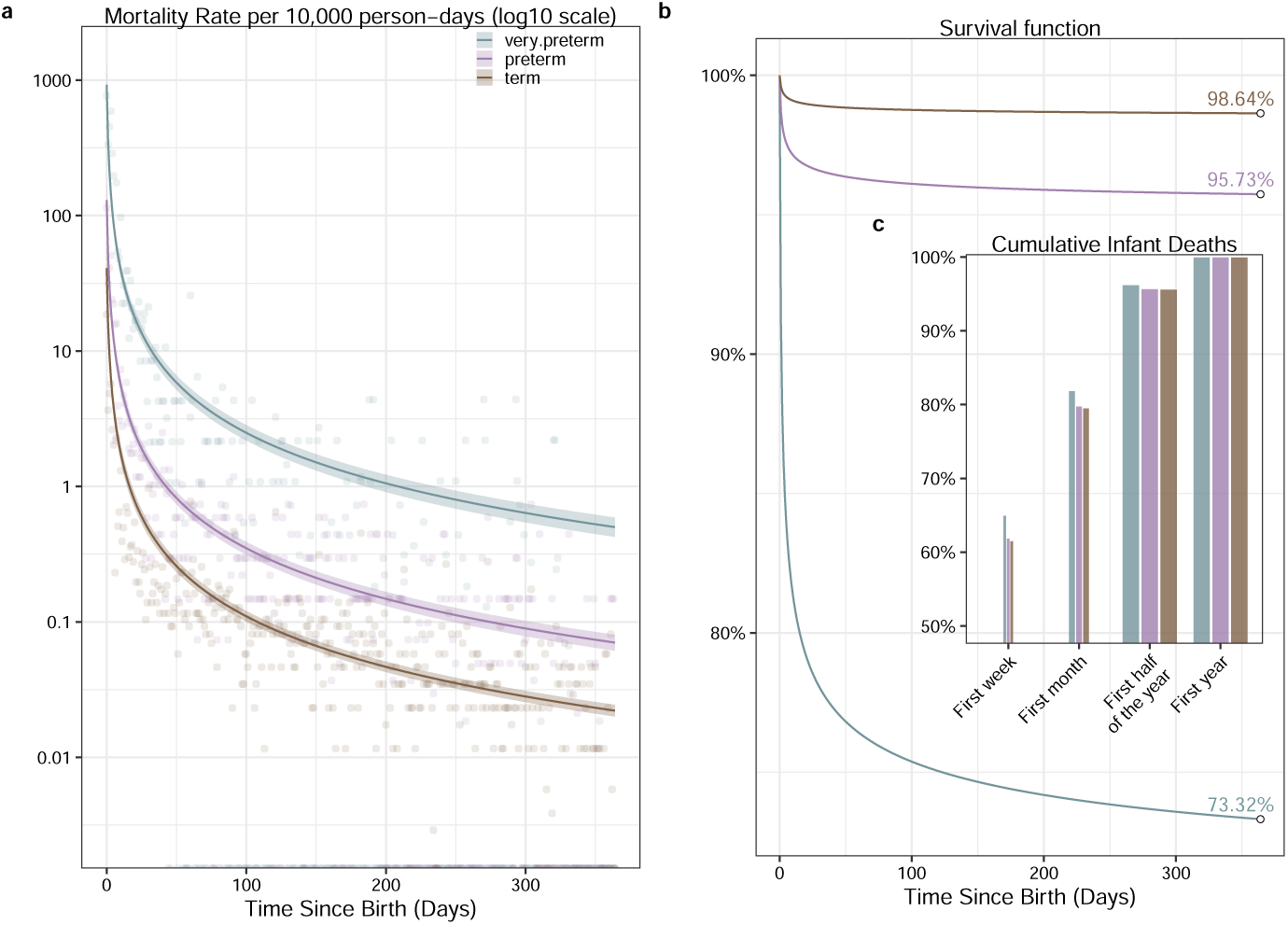
The infant mortality and survival function trends in the first year by gestational age groups since birth. **(a).** The estimated infant mortality rate per 10,000 person days with the base-10 log transformation in the first year since birth (x-axis) with the fitted model log D̂*_x_* = *α −* 1.25 *×* log(*x* + 0.88) +log(E*_x_*), with the fix effects *α* for Very preterm ([28, 32) weeks), Preterm ([32, 37) weeks), and Term ([37, 42) weeks) of −2.54, −4.50 and −5.66, respectively. The posterior median values of mortality rates are shown in line with 95% Cr.I. in ribbons. The corresponding Cr.I. are shown in Table 3. The empirical mortality rates are shown in dots. **(b).** and **(c).** showed the same estimates based on the posterior median value of mortality rates by gestational age groups (colour), with the same notations as in Fig. 1. The detailed information in **(c)**. is shown in Table 1.

**Table 1:**
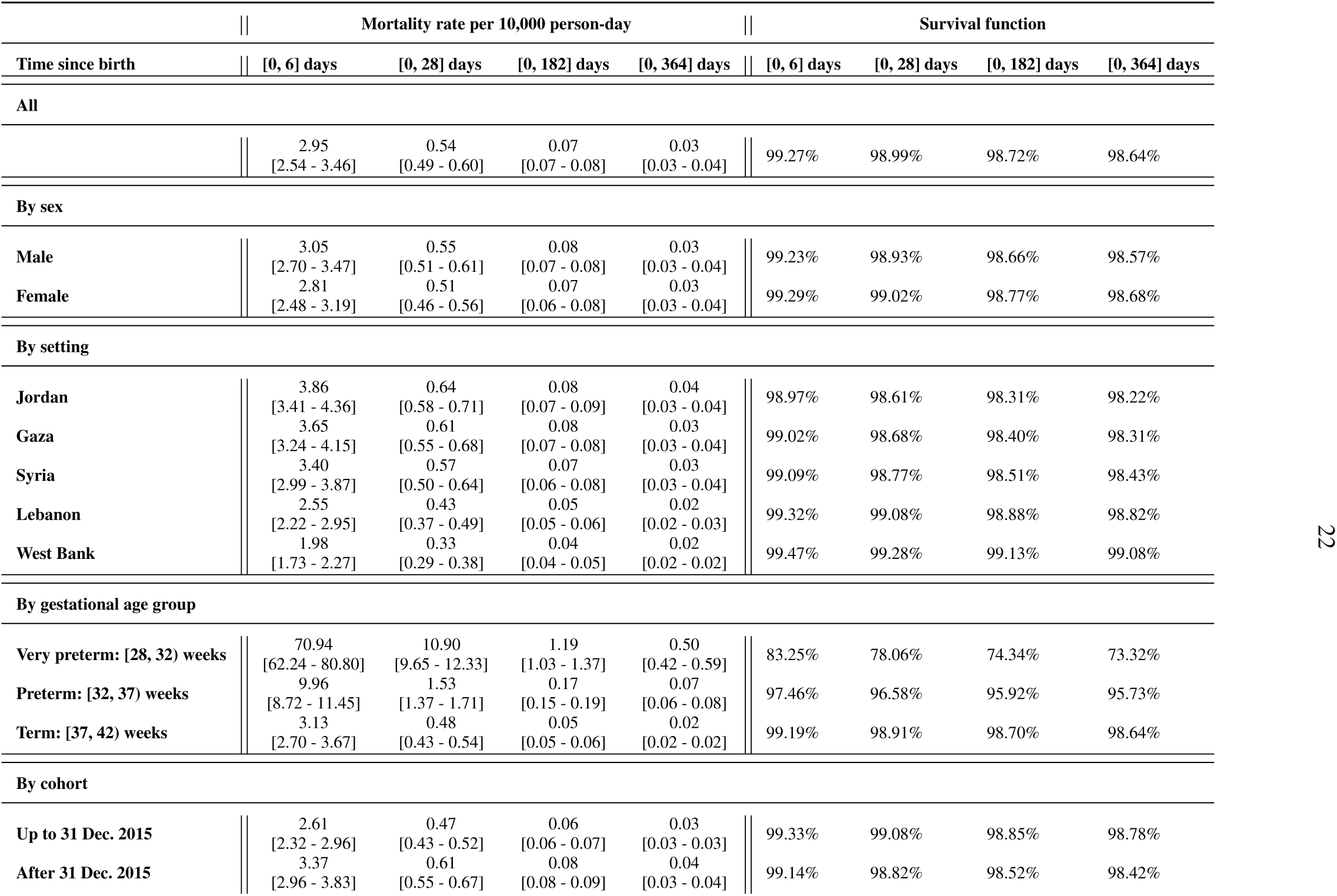
Summary table of the mortality rate and survival rate in each scenario.

## Results

### Age trajectories of infant mortality among Palestinian refugees

Fig. 1 shows daily mortality rates per 10,000 person-days of exposure (Fig. 1 **a**), the survival function over the first year of life (Fig. 1 **b**), and the cumulative age-at-death distribution of infant deaths (Fig. 1 **c** and Table 2) for Palestinian refugees in the full sample. Mortality risk declined sharply with age, with a rate of 2.95 per 10,000 person-days (95% Credible Interval [2.54, 3.46]) during the first week, decreasing to 0.54 [0.49, 0.60] over the first month (Table 1). Survival to one year of age was 98.64%, corresponding to a probability of dying within the first year of 13.6 per cent. Cumulatively, 55.46% of infant deaths in the first year of life occurred during the first week, and 73.80% had occurred by the end of the first month (Table 2).

**Table 2:** Summary table of cumulative age-at-death distribution of infant deaths in each scenario.

| Time since birth | [0, 6] days | [0, 28] days | [0, 182] days | [0, 364] days |
| --- | --- | --- | --- | --- |
| **All** |  |  |  |  |
|  | 55.46% | 73.80% | 93.63% | 100% |
| **By sex** |  |  |  |  |
| Male | 56.13% | 74.21% | 93.73% | 100% |
| Female | 56.13% | 74.21% | 93.73% | 100% |
| **By setting** |  |  |  |  |
| Jordan | 60.02% | 78.28% | 94.85% | 100% |
| Gaza | 60.00% | 78.26% | 94.84% | 100% |
| Syria | 59.97% | 78.24% | 94.84% | 100% |
| Lebanon | 59.93% | 78.21% | 94.82% | 100% |
| West Bank | 59.89% | 78.18% | 94.82% | 100% |
| **By gestational age group** |  |  |  |  |
| Very preterm: [28, 32) weeks | 64.98% | 82.53% | 96.21% | 100% |
| Preterm: [32, 37) weeks | 61.87% | 80.49% | 95.68% | 100% |
| Term: [37, 42) weeks | 61.53% | 80.25% | 95.62% | 100% |
| **By cohort** |  |  |  |  |
| Up to 31 Dec. 2015 | 56.52% | 74.54% | 93.84% | 100% |
| After 31 Dec. 2015 | 56.55% | 74.56% | 93.85% | 100% |

### Sex differences of infant mortality

We observe similar mortality profiles for males and females (Fig. 2). Mortality rates during the first week were 3.05 [2.70, 3.47] per 10,000 person-days for males and 2.81 [2.48, 3.19] for females (Table 1). Females exhibited a slight survival advantage over the first year of life (98.68% vs 98.57%). Differences in age-specific trajectories were minimal but systematic, corresponding to a modest female advantage.

### Variation by setting

Fig. 3 shows age-specific trajectories of infant mortality for Jordan, Gaza, Syria, Lebanon and the West Bank. Mortality rates followed similar age patterns (panel **a**) but differed markedly in level. The highest mortality rates were observed in Jordan and Gaza, while the lowest was in the West Bank. Across settings, the proportion of deaths occurring in the first week ranged from 59.89% to 60.02%, and by the first month, over 78.18% had occurred in all settings (Table 2). Survivorship over the first year of life reflected differences across settings (Table 1). In the West Bank, 99.08% of infants survived the first year of life, whereas in Jordan, Gaza, and Syria, survivorship was lower, ranging from 98.22% to 98.43%. This corresponds to the probability of dying within the first year, ranging from less than 1% in the West Bank to 1.78% in Jordan (Fig. 3 **b**), indicating a nearly twofold difference in the probability of death between the highest- and lowest-mortality settings.

### Preterm birth and high mortality

To understand how the timing of birth affects the infant mortality rate, we separated the observations into three gestational age groups (very preterm: [28, 32) weeks, preterm: [32, 37) weeks and term: [37, 42) weeks) (Fig. 4 **a**).

Mortality rates during the first week were 70.94 [62.24, 80.80] per 10,000 person-days among very preterm births, 9.96 [8.79, 11.45] among preterm births and 3.13 [2.70, 3.67] for term births (Table 1), indicating substantial variation across gestational age groups in the first week since birth. Mortality rates during the first month were 10.90 [9.65, 12.33] per 10,000 person-days among very preterm births, which was 22.7 times higher than in the term births group (0.48 [0.43, 0.54]). This differences remain over the first year, the mortality rate ratio between the very preterm and term groups increased to 25 (0.5 per 10,000 person-days in very preterm births versus 0.02 in term births).

Very preterm and preterm births exhibited a marked survival disadvantage over the first year of life (73.32% and 95.73%, respectively versus 98.64% among term births) (Fig. 4 **b** and Table 1). Cumulatively, 64.98% and 61.87% of deaths in the first year of life among very preterm and preterm births, respectively, occurred during the first week, and 82.53% (very preterm) and 80.49% (preterm) had occurred by the end of the first month. These patterns imply a higher concentration of early deaths compared with term births (61.53% during the first week and 80.25% in the first month) (Fig. 4 **c** and Table 2).

### Cohort differences

We separated the data into two cohorts born before (inclusive) and after 31 Dec 2015, respectively. Results in Fig. 5 show patterns of mortality relatively stable over cohorts, with a slight disadvantage for those born in the youngest cohort (Fig. 5 **a**).

**Fig. 5:**
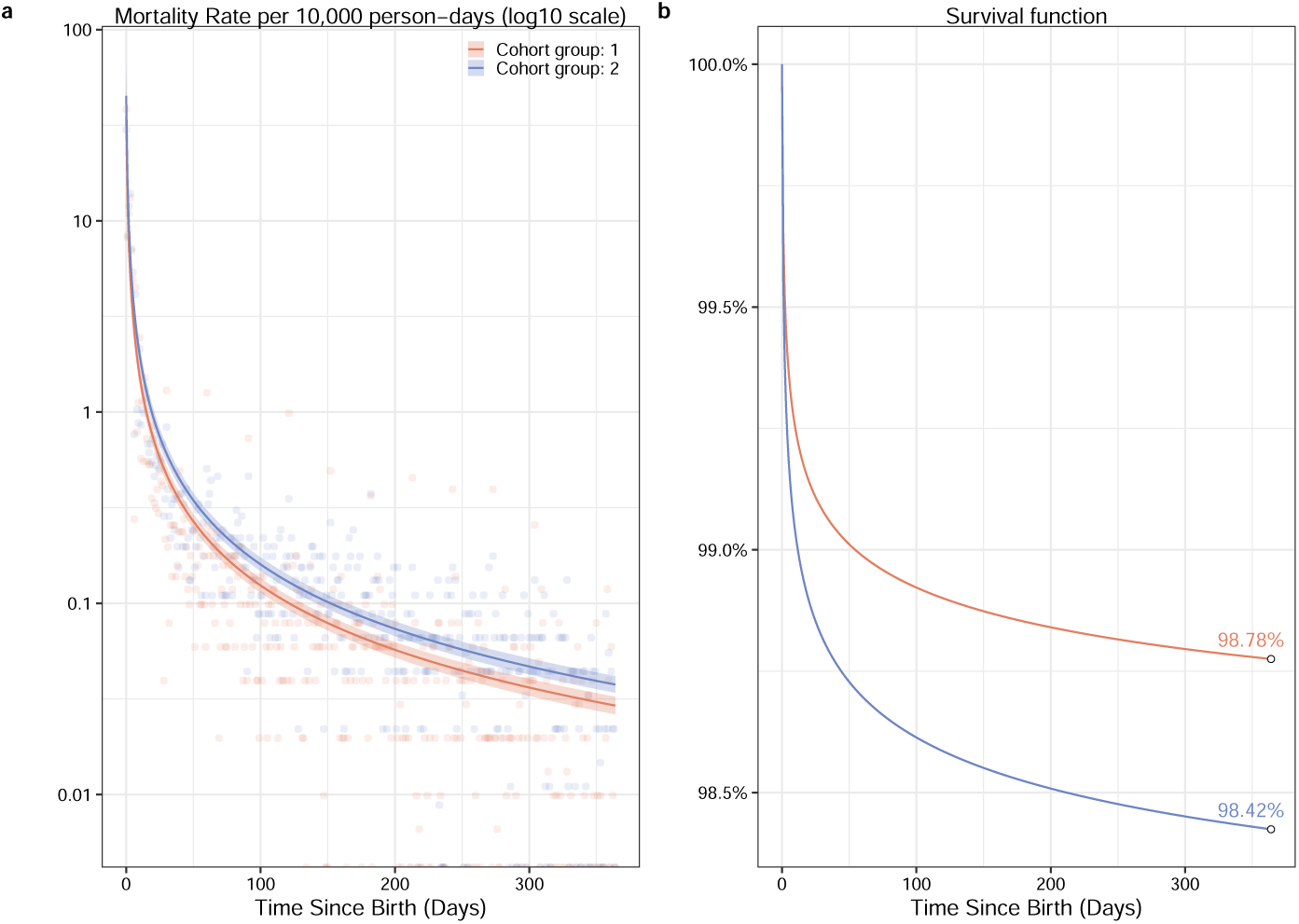
The infant mortality and survival function trends in the first year by cohort since birth. **(a).** The estimated infant mortality rate per 10,000 person days with the base-10 log transformation in the first year since birth (x-axis) with the fitted model log D̂*_x_* = *α−*1.12*×*log(*x*+0.66)+log(E*_x_*), with the fix effects *α* for children born up to 31 Dec 2015 (cohort group 1) of −6.12 and children born after 31 Dec 2015 (cohort group 2) of −5.86. The corresponding Cr.I. are shown in Table 3. The posterior median values of mortality rates are shown in line with 95% Cr.I. in ribbons. The empirical mortality rates are shown in dots. **(b).** showed the same estimates based on the posterior median value of mortality rates by cohort (colour).

**Table 3:**
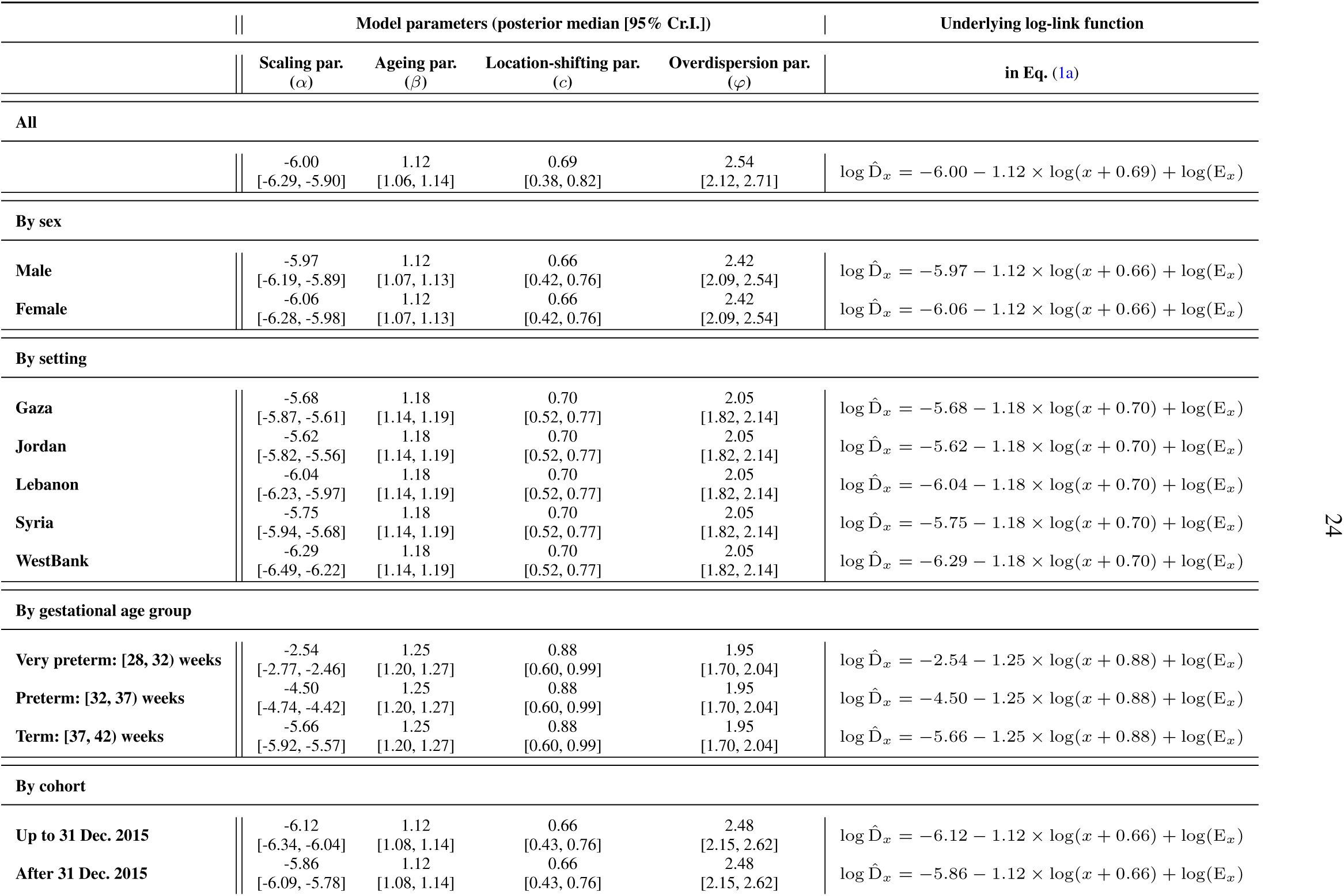
Summary table of the inferred model parameters with posterior median and 95% Credible intervals (Cr.I.)

Mortality rates during the first week were 2.61 [2.32, 2.96] per 10,000 person-days for infants born up to 31 Dec. 2015 and 3.37 [2.96, 3.83] after 31 Dec. 2015 (Table 1). Mortality rates during the first half year were 0.06 [0.06, 0.07] per 10,000 person-days among infants born up to 31 Dec. 2015, and 0.08 [0.08, 0.09] in the other group. In the first year since birth, the mortality rates were similar (0.03 per 10,000 person-days in group 1 versus 0.04 in group 2) (Table 1).

The recent 5-year cohort (group 2) exhibited a slight survival disadvantage over the first year of life (98.42% versus 98.78% among births up to 31 Dec. 2015) (Fig. 5 **b** and Table 1), indicating that differences in age-specific trajectories were minimal but systematic. As a sensitivity check in the recent 5-year cohort, we removed data in 2020 to exclude the potential impacts of the COVID-19 pandemic for a sensitivity check. We found no systematic difference in whether or not we included births in 2020 (Supplementary Tables 5–6).

## Discussion

### Summary of findings

Drawing on a unique electronic data linkage, we developed a statistical and demographic strategy that enabled reliable estimation of infant mortality trajectories, day-specific mortality hazards, age-at-death distributions and survivorship. Our results show variation in survival to the first year of life among Palestinian refugees across five settings, by sex, by gestational age group (very preterm, preterm and term), and by birth cohorts (2010-2015; 2016-2020). According to our estimates, 98.64% of infants survived to age 1, corresponding to infant mortality rate of 13.6 per 1000 live births. More than half of these deaths occurred within the first week of life, and a cumulative 70% by the end of the first month. We further estimated infant mortality trajectories across different sub-populations. Mortality profiles were similar by sex, with females showing a slight survival advantage (98.68% and 98.57%, respectively). By setting, instead, we observe moderate differences. Jordan, Gaza and Syria had the lowest survival (98.22%, 98.31% and 98.43%), while the West Bank had the highest (99.08%).However, the 95% credible intervals for infant mortality rates in Jordan, Gaza, and Syria overlap, suggesting no meaningful differences between these settings. When restricting to births occurring from the 28th gestational week onwards, infants born very preterm ([28, 32) weeks) and preterm ([32, 37) weeks) showed significantly lower survival rates compared to term births (73.32%, 95.73% vs. 98.64%), as expected [20]. Finally, we estimated differences in survival to age 1 across cohorts, and found that children born until 31 December 2015 had marginally higher infant survival than those born after (98.78% vs. 98.42%).

### Interpretation

Our findings on age-specific infant mortality are consistent with the broader literature on neonatal and infant mortality. The concentration of deaths in the first week of life is well-documented across diverse settings [21, 22], as well as the role of preterm birth in driving early mortality [23]. This age trajectory is also in line with previous studies conducted among Palestinian refugees, which estimated that between 59% and 74% of infant deaths occur within the first month of life [9]. The female survival advantage observed in our estimates is well-established across different populations and settings [24]. To facilitate comparison with the UNRWA preceding-birth estimates, we can express our cohort-based probability of dying as deaths before age 1 per 1,000 live births. This quantity is conceptually comparable to the infant mortality rate reported in previous studies [11], although our estimate is derived directly from cohort survival data, whereas their estimate is based in preceding birth techniques that reflects a retrospective survey-based approximation to infant mortality. In this sense, our estimate of 13.6 per 1,000 live births for the whole 2010-2020 period is slightly lower than the 18 per 1,000 reported in the most recent comparable study of Palestinian refugees across different settings for 2011 [11]. Looking at our estimates by birth cohort, later cohorts have slightly higher mortality rate (16 per 1,000 live births) than earlier ones (12 per 1,000 live births), which is consistent with the stalled or even worsening trends in infant mortality previously documented in the Palestinian refugee population [8]. These comparisons should be interpreted with caution, as data collection methods are not strictly comparable across studies.

The differences we observe in infant mortality across settings are broadly consistent with previous work. The only prior study to examine infant mortality among Palestinian refugees across five settings employed preceding birth method and found lower infant mortality among Palestinian refugees residing in the West Bank and Lebanon compared to other settings in 2008, similar to our results [9]. That study ranks infant mortality lowest in Syria, followed by Jordan, while in our estimates the ranking shifts, with Jordan showing the lowest survival, followed by Gaza and Syria. This change in ranking may partly reflect the impact of the three major Israeli military attacks in Gaza in 2008–2009, 2012, and 2014, which resulted in substantial civilian casualties and injuries [25]. Exposure to armed conflict can also indirectly increase infant mortality through disruptions to healthcare services including perinatal care, compromised food security, and worsening infectious disease, malnutrition, and sanitation conditions [26]. During this period, the Israeli blockade on Gaza intensified significantly, which limited the reconstruction of infrastructure damaged by armed conflict [27], disrupted efforts to train health professionals, and reduced both the volume and range of medical supplies able to cross checkpoints [26].Together, these factors may have exacerbated infant deaths in Gaza over the study period. It should be noted, however, that in both our estimates and prior work, the credible or confidence intervals across these three settings overlap.

To contextualise our results, we compared our survival to age 1 estimates to host country populations and showed that infant survival levels are very similar between Palestinian refugees and host country populations, particularly in Lebanon, Jordan, and Gaza (see Supplementary Table 7). In Syria, while we estimated higher survival among refugees than in the local population (98.43% vs. 98.05%), other studies found the opposite in 2008, with host country figures more favourable [9]. This shift is likely attributable to the Syrian civil war, which began in 2011 and had severe effects on fetal, neonatal, and infant mortality in the general population [28]. The West Bank stands out with comparatively higher infant survival relative to the State of Palestine average, consistent with the findings of prior work [9]. This advantage is commonly attributed to the specialised and consistent primary healthcare provided by UNRWA including perinatal care and immunisation programmes which often exceeds the services available to non-refugee populations [7]. The achievements of UNRWA include early initiation of antenatal care, promotion of breastfeeding and near-universal vaccination coverage, which have improved neonatal and infant survival considerably [29].

### Limitations

While the data used in this study are unique in providing detailed information on infant mortality trajectories across five settings, they have several limitations. First, there are issues related to delays in death registration, data completeness, and reporting. This may also mean that some very early neonatal deaths were misreported as stillbirths. We mitigate these issues through a harmonisation and smoothing approach applied to hazard trajectories, combined with an imputation procedure that has been shown to produce reliable mortality estimates from highly granular data in other contexts [15, 16]. Second, the sample results in fewer observations, as we stratified the sample more granularly (e.g. sex and field) which prevented more detailed analyses such as stratification by clinic. Nonetheless, our models appeared robust and performed well across all specifications used. Third, access to UNRWA services varies considerably by setting, and not all refugees use them and the socioeconomic profile of UNRWA service users can differ from one setting to another [30]. Importantly, while our sample is restricted to UNRWA service users, this does not imply that all infant deaths are recorded in clinical settings. Even among UNRWA users, deaths can occur outside of facilities, especially during periods of armed conflict, when births may take place in unsafe environments with a very high risk of intrapartum complications and neonatal death [31]. Taken together, these issues are most likely to result in an underestimation of mortality rates, and our results should therefore be interpreted as lower-bound estimates.

### Conclusion

Our estimates derived from electronic health records of Palestinian refugees show broadly consistent results with previous estimates produced using survey data. This suggests that, in settings where well-established routine health information systems exist, electronic health records data can complement traditional data collection approaches. Our results further show that, despite the limitations of routinely collected data (e.g. incomplete records and selection on healthcare users), robust and comparable estimates can be obtained through Bayesian methods, which are well-suited to handling such data challenges. Where such systems are already in place, electronic health records also offer additional advantages over traditional survey methods, as they draw on larger samples, provide real-time estimates, and are less costly to implement [4]. These advantages become particularly consequential in forced displacement settings, as conflict and resource constraints frequently disrupt survey fieldwork. [3].

## Conclusion

Drawing on a unique electronic data linkage, this study examined age-specific trajectories of infant mortality during the first year of life among Palestinian refugees stratified by sex, gestational age, and setting. Our findings reveal broadly consistent mortality trajectories by sex and gestational age with prior work, while also identifying meaningful differences across refugee settings. Overall, our findings highlight the potential of electronic health records for mortality monitoring in forced displacement settings.

## Data Availability

The data used in the present study is not publicly available. The replication files for this paper include customised functionality written in R (version 4.5.0). The code, and all harmonised output data pertaining to our analysis, is hosted on GitHub https://github.com/selinkoksal/inf-mortality-palestine

## Ethics approval

This project is conducted as part of a broader study (ESRC Grant number: ES/Z503265/1) that received ethical approval from the London School of Hygiene & Tropical Medicine (LSHTM) Research Ethics Committee (reference 25467, last updated 18/11/2024)

## Contributions

Conceptualization: JMA, OC; Data curation: SK, ZJ, SA; Formal analysis: JMA, SK, YC; Methodology: JMA, YC; Software: JMA, SK, YC; Visualization: JMA, YC; Project administration: JMA, OC; Supervision: JMA, OC; Writing – original draft: JMA, SK, YC; Writing – review & editing: All authors.

## Data and materials availability

The replication files for this paper include customised functionality written in R (version 4.5.0). The code, and all harmonised output data pertaining to our analysis, is hosted on GitHub https://github.com/selinkoksal/inf-mortality-palestine.

## Acknowledgements and funding

Wellcome Trust CDA grant 07859/Z/23/Z (SK, YC, JMA). The Economic and Social Research Council [Grant number: ES/Z503265/1] (OC, ZJ). We used AI-assisted tools for proofreading purposes.

## Patient and Public Involvement statement

Patients and the public were not involved in the design, conduct, reporting, or dissemination of this research.

## References

[1] Nott J, Kaunda-Khangamwa B, Mathanga DP, Molenaar J, Taithe B, Phiri C, et al. Beyond the Demographic and Health Survey: on the past and future of population health surveillance. BMJ Global Health. 2026;11(2):e022023.

[2] Levy BS, Sidel VW. Documenting the effects of armed conflict on population health. Annual review of public health. 2016;37(1):205–18.

[3] Calvert C, Kaushal S, Banke-Thomas A, Jamaluddine Z, Matovu B, Riches J, et al. Rising to the challenge: lessons learnt from the Global Women’s Research Society (GLOW) conference for women’s and newborn health in the context of global crises. Sexual and reproductive health matters. 2025;33(1):2525656.

[4] Casey JA, Schwartz BS, Stewart WF, Adler NE. Using electronic health records for population health research: a review of methods and applications. Annual review of public health. 2016;37(1):61–81.

[5] Sabatinelli G, Pace-Shanklin S, Riccardo F, Shahin Y. Palestinian refugees outside the occupied Palestinian territory. The Lancet. 2009;373(9669):1063–5.

[6] Jamaluddine Z, Seita A, Ballout G, Al-Fudoli H, Paolucci G, Albaik S, et al. Establishment of a birth-to-education cohort of 1 million Palestinian refugees using electronic medical records and electronic education records. International journal of population data science. 2023;8(1):2156.

[7] Khawaja M. The extraordinary decline of infant and childhood mortality among Palestinian refugees. Social Science & Medicine. 2004;58(3):463–70.

[8] van den Berg MM, Khader A, Hababeh M, Zeidan W, Pivetta S, Abd El-Kader M, et al. Stalled decline in infant mortality among Palestine refugees in the Gaza Strip since 2006. PloS one. 2018;13(6):e0197314.

[9] Riccardo F, Khader A, Sabatinelli G. Low infant mortality among Palestine refugees despite the odds. Bulletin of the World Health Organization. 2011;89(4):304–11.

[10] van den Berg MM, Madi HH, Khader A, Hababeh M, Zeidan W, Wesley H, et al. Increasing neonatal mortality among Palestine refugees in the Gaza Strip. PloS one. 2015;10(8):e0135092.

[11] van den Berg MM, Khader A, Hababeh M, Zeidan W, Seita A. Infant and neonatal mortality among Palestine refugees in Gaza, West Bank, Lebanon, and Jordan: an observational study. The Lancet. 2017;390:S10.

[12] Kozuki N, Walker N. Exploring the association between short/long preceding birth intervals and child mortality: using reference birth interval children of the same mother as comparison. BMC public health. 2013;13(Suppl 3):S6.

[13] United Nations High Commissioner for Refugee(UNHCR). Figures at a Glance; 2025. Accessed on 16 Feb, 2026. Available from https://www.unhcr.org/about-unhcr/overview/figures-glance.

[14] Jamaluddine Z, Paolucci G, Ballout G, Al-Fudoli H, Day LT, Seita A, et al. Classifying caesarean section to understand rising rates among Palestinian refugees: results from 290,047 electronic medical records across five settings. BMC pregnancy and childbirth. 2022;22(1):935.

[15] Schöley J. The Dynamics of Ontogenescence: Modelling Age Trajectories of Feto-Infant Mortality. Syddansk Universitet; 2020. https://osf.io/cfypa/files/2ksyp.

[16] Schöley J. The impact of population heterogeneity on the age trajectory of neonatal mortality. Demographic Research. 2025;53:187–218.

[17] Colchero F, Aburto JM, Archie EA, Boesch C, Breuer T, Campos FA, et al. The long lives of primates and the ‘invariant rate of ageing’hypothesis. Nature communications. 2021;12(1):3666.

[18] Stan Development Team and their assignees. Gaussian Processes; 2024. Accessed Oct. 10, 2024. https://mc-stan.org/docs/stan-users-guide/gaussian-processes.html#fit-gp.section.

[19] Bürkner PC. brms: An R package for Bayesian multilevel models using Stan. Journal of statistical software. 2017;80:1–28.

[20] Cao G, Liu J, Liu M. Global, regional, and national incidence and mortality of neonatal preterm birth, 1990-2019. JAMA pediatrics. 2022;176(8):787–96.

[21] Hug L, Alexander M, You D, Alkema L. National, regional, and global levels and trends in neonatal mortality between 1990 and 2017, with scenario-based projections to 2030: a systematic analysis. The Lancet Global Health. 2019;7(6):e710–20.

[22] Sankar M, Natarajan C, Das R, Agarwal R, Chandrasekaran A, Paul V. When do new-borns die? A systematic review of timing of overall and cause-specific neonatal deaths in developing countries. Journal of perinatology. 2016;36(1):S1–S11.

[23] Oza S, Lawn JE, Hogan DR, Mathers C, Cousens SN. Neonatal cause-of-death estimates for the early and late neonatal periods for 194 countries: 2000–2013. Bulletin of the World Health Organization. 2014;93:19–28.

[24] Fuse K, Crenshaw EM. Gender imbalance in infant mortality: A cross-national study of social structure and female infanticide. Social Science & Medicine. 2006;62(2):360–74.

[25] Mosleh M, Dalal K, Aljeesh Y, Svanström L. The burden of war-injury in the Palestinian health care sector in Gaza Strip. BMC international health and human rights. 2018;18(1):28.

[26] Leone T, Alburez-Gutierrez D, Ghandour R, Coast E, Giacaman R. Maternal and child access to care and intensity of conflict in the occupied Palestinian territory: a pseudo-longitudinal analysis (2000–2014). Conflict and health. 2019;13(1):36.

[27] Manduca P, Al Baraquni N, Al Baraquni L, Abadi DA, Abdallah H, Hamad GA, et al. Hospital centered surveillance of births in Gaza, Palestine, 2011–2017 and heavy metal contamination of the mothers reveals long-term impact of wars. Reproductive Toxicology. 2019;86:23–32.

[28] DeJong J, Ghattas H, Bashour H, Mourtada R, Akik C, Reese-Masterson A. Reproductive, maternal, neonatal and child health in conflict: a case study on Syria using Countdown indicators. BMJ global health. 2017;2(3).

[29] United Nations Relief and Works Agency for Palestine Refugees in the Near East. Annual Report 2022. UNRWA; 2023. Available from: https://www.unrwa.org/sites/default/files/content/resources/annual_report_2022_final_version_compressed-july_2023.pdf.

[30] Lapeyre F, Al Husseini JJ, Bocco R, Brunner M, Zureik E. The living conditions of the Palestine refugees registered with UNRWA in Jordan, Lebanon, the Syrian Arab republic, the Gaza Strip and the West Bank. Lebanon, the Syrian Arab Republic, the Gaza Strip and the West Bank (May 16, 2011). 2011.

[31] Wick L, Hassan S. No safe place for childbirth: women and midwives bearing witness, Gaza 2008–09. Reproductive health matters. 2012;20(40):7–15.

