## Supplementary Materials for "Age trajectories of infant mortality among Palestinian refugees born in 2010–2020: evidence from electronic health records"

**4 Supplementary Figures** **12**

### 1 Previous infant mortality estimates among Palestinian refugees

Supplementary Table 1: Infant mortality rate estimates for Palestine refugees by setting and year

| Population | Year(s) | Data source | Reference | IMR (per 1,000) |
|---|---|---|---|---|
| Gaza | 1960 | UNRWA | (van den Berg et al. 2018) | 127.0 |
| | 1967 | UNRWA | (van den Berg et al. 2018) | 82.0 |
| | 1995 | UNRWA | (Riccardo et al. 2011) | 33.0 |
| | 2001 | UNRWA | (Riccardo et al. 2011) | 25.2 |
| | 2006 | UNRWA | (Riccardo et al. 2011) | 20.2 |
| | 2011 | UNRWA | (van den Berg et al. 2015) | 22.4 |
| | 2013 | UNRWA | (van den Berg et al. 2018) | 22.7 |
| | 2019 | MICS | (UNICEF State of Palestine 2021) | 17.0 |
| Syria | 1995 | UNRWA | (Riccardo et al. 2011) | 29.0 |
| | 2001 | UNRWA | (Riccardo et al. 2011) | 28.2 |
| | 2006 | UNRWA | (Riccardo et al. 2011) | 28.2 |
| Jordan | 1995 | UNRWA | (Riccardo et al. 2011) | 32 |
| | 1999 | Jordan refugee camps survey | (Khawaja 2004) | 25.0 |
| | 2001 | UNRWA | (Riccardo et al. 2011) | 22.5 |
| | 2006 | UNRWA | (Riccardo et al. 2011) | 22.6 |
| Lebanon | 1995 | UNRWA | (Riccardo et al. 2011) | 35 |
| | 1999 | Lebanon refugee camps survey | (Khawaja 2004) | 32.0 |
| | 2006 | UNRWA | (Riccardo et al. 2011) | 19.0 |
| | 2011 | UNRWA | (Riccardo et al. 2011) | 19.2 |
| West Bank | 1995 | West Bank and Gaza Strip Demographic Survey | (Khawaja 2004) | 25.0 |
| | 2001 | UNRWA | (Riccardo et al. 2011) | 15.3 |
| | 2006 | UNRWA | (Riccardo et al. 2011) | 19.5 |
| | 2014 | MICS | (van den Berg et al. 2018) | 17.0 |

### 2 Data

| Sample | Births | Deaths † |
|---|---|---|
| **Full** | 972 743 | 14 009 |
| *Sex** | | |
| Male | 502 413 | 6 964 |
| Female | 468 721 | 5 436 |
| *Field* | | |
| Gaza | 438 134 | 6 320 |
| Jordan | 267 624 | 4 442 |
| Lebanon | 56 075 | 740 |
| Syria | 64 093 | 1 043 |
| West Bank | 146 817 | 1 464 |
| *Gestational age* | | |
| Very preterm | 6 684 | 2 158 |
| Preterm | 70 544 | 3 155 |
| Term | 886 582 | 7 681 |
| *Cohort* | | |
| Up to 2015 | 512 914 | 7 248 |
| 2016 onwards | 459 829 | 6 761 |

† These figures show all deaths recorded in the sample, including those occurring after age 1. Statistical analyses were restricted to deaths within the first year of life, as the objective of this paper is to estimate infant mortality.

* There are 1,609 missing values on the sex variable; these are imputed for the main analyses but shown as missing here, as this table presents raw data.

Supplementary Table 2: Number of births and deaths by subpopulations used in the analyses

#### 2.1 Data quality checks

To assess the quality of mortality data, we used two indicators.First, we calculated the ratio of very early neonatal deaths (0–1 completed days) to early neonatal deaths (2–6 completed days), a common indicator for assessing the quality of neonatal mortality (Ali et al. 2023). In this indicator, values greater than 2.4 are considered plausible for high-quality data. Lower values may indicate displacement of deaths from the first two days of life into later days. This means under-reporting of deaths at 0–1 days, or over-reporting at 2–6 days or misreporting of very early live births as stillbirths. Then, we calculated heaping ratios at selected focal points: days 7, 10, 15, 30, 60, and every 30 days through day 330, corresponding to 11 months. These focal points were chosen to capture heaping at round numbers, the end of the first week, and subsequent monthly intervals. The heaping ratio at each focal point is calculated as the number of deaths occurring on that day divided by the total number of deaths occurring within a ±2-day window centred on the focal point, present in Equation (1) below:

$$H(d) = \frac{D_d}{\left(\sum_{k=d-2}^{d+2} D_k\right)/5} \tag{1}$$

where $D_d$ is the number of deaths occurred at day d. $H(d) \approx 1$ indicates no heaping, $H(d) > 1$ indicates excess reporting at day $d$. Values of $H(d)$ between 1.2 and 1.5 indicate mild heaping, while $H(d) > 1.5$ indicates severe heaping. The ratio of very early deaths to early deaths suggested as a quality check indicator by Ali et al. (2023), where values over 2.4 is considered as plausible. Supplementary Table 3 documents the results of the quality assessment for those deaths for which there is full age-at-death information. The ratio of very early deaths to early deaths is 0.88, well below the reference value of 2.4, suggesting potential under-reporting at 0–1 days and/or displacement to later days within the early neonatal period. We also observe strong heaping at day 7 (H = 2.26), indicating a strong concentration of reported deaths at the end of the first week of life. Additional strong heaping is observed on days 10 (H = 1.87), 30 (H = 2.80), 120 (H = 3.02), 240 (H = 2.93) and 270 (H = 2.00), suggesting an over-representation of deaths at approximately months 1, 4, 8 and 9. Mild heaping was present at day 15 (H = 1.16) and day 300 (H = 1.36). In contrast, days 60 (H = 0.47), 90 (H = 0.22), 150 (H = 0.38), and 180 (H = 0.52) showed heaping ratios below 1, indicating relative under-representation compared with adjacent days

| Indicator | Heaping Index |
|---|---|
| Very early (0–1 days) / Early (2–6 days) | 0.88 |
| Heaping at day 7 | 2.26 |
| Heaping at day 10 | 1.87 |
| Heaping at day 15 | 1.16 |
| Heaping at day 30 | 2.80 |
| Heaping at day 60 | 0.47 |
| Heaping at day 90 | 0.22 |
| Heaping at day 120 | 3.02 |
| Heaping at day 150 | 0.38 |
| Heaping at day 180 | 0.52 |
| Heaping at day 210 | 1.56 |
| Heaping at day 240 | 2.93 |
| Heaping at day 270 | 2 |
| Heaping at day 300 | 1.36 |
| Heaping at day 330 | 0 |

Supplementary Table 3: Data quality assessment indicators for full sample

### 3 Statistical modelling

In this section, we provide more details about the statistical framework setting to model infant mortality rates.

#### 3.1 Full model specification

We aligned with the general shift power function to infer the mortality rates, with the hazard rate formula of

$$h(x) = \alpha(x + c)^{\beta} e^{-px}.$$

In the age domain $x$, we set $p = 0$ to simplify the model, because the exponential term $e^{-px}$ controlled the hazard growth among adults, and we focused only on infant mortality estimation here. We introduced the location-shifting parameter $c$, which quantifies the threshold age at which the mortality process begins. We identified the value of $c$ in the modelling process because the inferred $c$ could differ across model scenarios. Parameter $\alpha$ is the scaling effect, and parameter $\beta$ represents the ageing effects. In the infant mortality modelling, $\beta$ is negative to decrease the power law. We analysed mortality counts using this parametric model developed by Jonas that incorporates demographic theory (Schöley 2020) modelling mortality risk as a power-law. We extended such method in a Bayesian state-of-the-art framework to incorporate demographic structure into the mortality shape. The deterministic model also reduced the extent of age heaping (main text).

As an extension of the model setting in the method section, we presented the full model specification with prior distributions below.

$$\begin{aligned}
\mathrm{D}_x^s &\sim \text{Negative Binomial}(\hat{\mathrm{D}}_x^s, \varphi) && \text{(2a)}\\
\log \hat{\mathrm{D}}_x^s &= \alpha^s + \beta \times \log(x^s + c) + \log \mathrm{E}_x^s && \text{(2b)}\\
\text{with weighted likelihood} & \\
\log L &= \sum_i \tilde{w}_i \log p(\mathrm{D}_x^s \mid \alpha^s, \beta, c, \varphi) && \text{(2c)}\\
w_x &= \mathbb{I}_{x<7} \times w_0 + \mathbb{I}_{x\geq 7} \times 1 && \text{(2d)}\\
\tilde{w}_x &= w_x / \text{mean}(w) && \text{(2e)}\\
\text{with priors} & \\
\alpha^s &\sim \text{Normal}(0, 5^2) && \text{(2f)}\\
\beta &\sim \text{Gamma}(5, 5) && \text{(2g)}\\
c &\sim \text{Gamma}(5, 5) && \text{(2h)}\\
\varphi &\sim \text{Exponential}(1) && \text{(2i)}
\end{aligned}$$

In Eq. (2a), $\hat{D}_x^s$ is the expected number of mortality counts of strata $s$ and on day $x$ since birth and $\varphi$ is the overdispersion parameter to capture the large variance from the observations.

We inferred the infant mortality rates with the developed Bayesian model, considering the death counts at the daily level since birth in the following four scenarios,

- overall level;
- stratified by sex;
- stratified by setting (Gaza, Jordan, Lebanon, Syria, and West Bank);
- stratified by gestational age ([28, 32) weeks as very preterm, [32, 37) weeks as preterm, and [37, 42) weeks as term);
- stratified by cohort (group 1: up to 31 Dec. 2015 and group 2: after 31 Dec. 2015).

#### 3.2 Computational implementation and model diagnostics

In each scenario, we implemented our developed Bayesian model using the Hamiltonian Monte Carlo algorithm in `brm` version 2.23.0 (Bürkner 2017) package with `rstan` version 2.32.7 in `Stan` version 2.32.2. Four chains in parallel, with 1,000 warm-ups and 4,000 sampling iterations each, converged and mixed well, confirmed by a maximum Gelman-Rubin diagnostic ($\hat{R}$) below 1.001 and a minimum effective sample size (Neff) above 2000. Trace plots in the overall scenario of four model parameters with the smallest Neff were shown in Supplementary Figure 2. The trace plots indicate that four chains per model parameter mix well and show no apparent non-stationary behaviour, consistent with convergence. Posterior predictive checks were performed by comparing observed outcomes with generated data from the posterior predictive distribution. More than 97% of observations were within 95% posterior predictive intervals (Supplementary Figures 3).

#### 3.3 Model comparison and sensitivity analyses

Below, we describe how we developed the central model and the types of model specifications we have assessed. We started with the assumption of likelihood and progressed to the assumption of the age trajectory. We showcased the model's forecasting performance to highlight and validate our assumption about the age patterns. Then, we compared our Bayesian regression framework to the Generalised Linear Model framework.

##### 3.3.1 Likelihood specification

Mortality counts were modelled by the Negative Binomial distribution to accommodate the overdispersion relative to the Poisson distribution assumption. We examined the ratio of variance to mean in each modelling stratum, and a ratio exceeding 100 in the overall modelling scenario motivated us to embed the overdispersion parameter in the model assumptions. In the other stratified scenarios, we observed even larger ratios, suggesting the need to include the overdispersion parameter.

In the preliminary analysis, we observed lower values of death counts in the first week relative to the observations, attributable to the highly concentrated early neonatal period, although the posterior predictive check was 97% (Supplementary Figure 4). We believed that death records during the early neonatal period should be high, so that the predicted data points in the early days after birth should be similar to or higher than the observed levels. To ensure that the model adequately captures the early mortality peak, we assigned greater weight to the likelihood for the first week. We experimented with weights of 2, 5, 10, 15, and 20 for the first 7 days and rescaled the weights to a mean of 1 so that the overall information about likelihood remained unchanged. We graphically compared the posterior predicted mortality count curves with the observations (Supplementary Figures 3–8). The posterior predicted mortality counts were generated from the posterior predictive distribution, accounting for the uncertainty in the inferred model parameters and the random noise from the Negative Binomial distribution. We found that with $w_0 = 5$, the median value of the predicted death counts on the first day since birth was close to the observation (Supplementary Figures 3), and this situation was not improved when we increased the weights (Supplementary Figures 6–8). Additionally, we analytically compared the model diagnostics ($\hat{R}$, effective sample size), Bayes $R^2$, LOO-CV ELPD with the associated standard error (ELPF SE), posterior predicted check (PPC), mean absolute error (MAE) and root mean square error (RMSE) (Supplementary Table 4). The model diagnostics were quite similar, suggesting well-mixed and converged results across all models. The LOO-CV ranking suggested that a weight of 5 was the best among models with weights greater than 5, but it was not significantly different from 0, given the large standard error. Regarding the MAE and RMSE, we preferred the model with $w_0 = 5$ as the central model.

| **Weight** | **Maximal $\hat{R}$** | **Minimal Neff** | **Bayes $R^2$** | **ELPD** | **ELPD SE** | **PPC** | **MAE** | **RMSE** |
|---|---|---|---|---|---|---|---|---|
| 1 | 1.00 | 4085 | 0.79 | -1172 | 30 | 97% | 15.02 | 78.40 |
| 2 | 1.00 | 3882 | 0.86 | -1205 | 42 | 97% | 13.87 | 64.95 |
| 5 | 1.00 | 3283 | 0.90 | -1292 | 91 | 97% | 13.36 | 58.94 |
| 10 | 1.00 | 3215 | 0.91 | -1418 | 168 | 97% | 13.59 | 57.92 |
| 15 | 1.00 | 3333 | 0.92 | -1524 | 235 | 97% | 13.86 | 57.89 |
| 20 | 1.00 | 3057 | 0.92 | -1616 | 293 | 97% | 13.98 | 58.26 |

Supplementary Table 4: Summary table of the model diagnostics and performance.

##### 3.3.2 Sensitivity analysis on age trajectory assumption

In the central analysis, we assumed that age-specific mortality rate patterns were identical across strata, i. e, the ageing parameter $\beta$ and the location-shifting parameter $c$ were shared across strata. By comparison, we inferred the age trajectories of mortality rates independently for each stratum, and forecasted for the following 200 days for visualisation. For simplicity, we compared the trends of the mortality rates side by side in Supplementary Figures 9–11.

Intuitively, in the sex-specific modelling case, we observed that before the first half year since birth, the age trajectories under the two assumptions are similar. However, the estimated male mortality rates within the individual modelling framework were lower than female rates after the first year since birth, which was not plausible in reality (Supplementary Figures 9). This difference can be explained by the inferred model parameters. In the individual modelling case, the ageing parameter of males was inferred as 1.15 [1.08, 1.17] with the location-shifting parameter of 0.72 [0.40, 0.87]. For females, the ageing parameter was slightly lower: 1.10 [1.03, 1.12], with a location-shifting parameter of 0.68 [0.38, 0.82].

In the setting scenario, we also observed overlapping age trajectories across settings under the independent modelling assumption (Supplementary Figure 10). In Supplementary Figures 11, we investigated the consistently higher mortality rates among very preterm births and lower rates among term births in the central analysis, compared to the results in independent modelling assumptions. However, after the first half year since birth, the gap across three age groups was decreasing in the independent modelling assumption.

In parallel, we also examined the assumption on the scaling parameter $\alpha^s$ when we have strata in the scenario. We introduced a shape parameter to flexibly control the variance of the prior distributions of $\alpha^s$, i. e, we changed the fixed effects to random effects. Besides the longer sampling time, the model performance and results were similar using fixed or random effects on $\alpha^s$. Therefore, we chose the simple model and kept the scaling parameter as the fixed effects.

###### 3.3.3 Framework comparison

Previous literature applied the Generalised Linear Model (GLM) to model the infant mortality rates. In this section, we compared our Bayesian framework with the GLM in the Frequentist framework.

Based on the previous Generalised Linear Model with Poisson assumption (Schöley 2020), we aligned with our assumption in the central model and modified the GLM model with Negative Binomial assumption. We applied the same initial weights in the first week as in the central model, but the estimated mortality rates were lower than the empiricals in the GLM setting, and required more weights to leverage the estimates in order to capture the early neonatal period (Supplementary Figure 16). To find the best-fitted value of the location-shifting parameter $c$, we used the 'optimize' function within the interval $[0.5, 1.5]$, with the initial value of 1. We fixed that value and inferred overdispersion for bootstrapping 1,000 times to propagate the uncertainty intervals.

Besides the relatively longer running time to sample the predicted estimates 1,000 times, we compared two models by MAE, RMSE, and coverage rate to validate the benefits of using the Bayesian framework. Estimates from the Bayesian model achieved lower MAE (13.36 vs 16.31) and RMSE (58.94 vs 69.53) compared with the frequentist estimator. The coverage rates were similar between the two approaches, i. e., around 97%. The Bayesian framework is flexible for further development in complex contexts, including the location-shifting parameter estimation within the sampling. The advantages of scalability, incorporation of prior knowledge and ease of uncertainty quantification prompted us to use the Bayesian model as our central analysis.

###### 3.3.4 Sensitivity analysis on removing data in 2020 for cohort-specific modelling

As we estimated slightly higher infant mortality rates in the cohort (2016-2020) than in the first six years (2010-2015), we assumed the 2020 observations would drive higher mortality rates due to the COVID-19 pandemic. We presented the comparison results in Supplementary Table 5 because it was difficult to directly investigate the differences from the figures. Here, we observed differences in mortality rates in the first week, and no substantial changes in the first month, first half-year, and first year since birth.

| | Mortality rate per 10,000 person-day | | | | Survival function | | | |
|---|---|---|---|---|---|---|---|---|
| **Time since birth** | **First week** | **First month** | **First half year** | **First year** | **First week** | **First month** | **First half year** | **First year** |
| **By cohort, including year 2020** | | | | | | | | |
| **Up to 31 Dec. 2015** | 2.61<br>[2.32 - 2.96] | 0.47<br>[0.43 - 0.52] | 0.06<br>[0.06 - 0.07] | 0.03<br>[0.03 - 0.03] | 99.33% | 99.08% | 98.85% | 98.78% |
| **After 31 Dec. 2015** | 3.37<br>[2.96 - 3.83] | 0.61<br>[0.55 - 0.67] | 0.08<br>[0.08 - 0.09] | 0.04<br>[0.03 - 0.04] | 99.14% | 98.82% | 98.52% | 98.42% |
| **By cohort, excluding year 2020** | | | | | | | | |
| **Up to 31 Dec. 2015** | 2.58<br>[2.27 - 2.94] | 0.47<br>[0.43 - 0.52] | 0.06<br>[0.06 - 0.07] | 0.03<br>[0.03 - 0.03] | 99.35% | 99.1% | 98.86% | 98.79% |
| **After 31 Dec. 2015** | 3.38<br>[2.96 - 3.87] | 0.62<br>[0.56 - 0.68] | 0.08<br>[0.08 - 0.09] | 0.04<br>[0.03 - 0.04] | 99.14% | 98.82% | 98.52% | 98.41% |

Supplementary Table 5: Summary table of the cohort-specific mortality rate and survival rate in each scenario.

Regarding the inferred model parameters in Supplementary Table 6, observations in 2020 not significantly affected the model estimations.

| | Model parameters (posterior median [95% Cr.I.]) | | | | Underlying log-link function |
|---|---|---|---|---|---|
| | **Scaling par.** ($\alpha$) | **Ageing par.** ($\beta$) | **Location-shifting par.** ($c$) | **Overdispersion par.** ($\varphi$) | **in Eq.** (2a) |
| **By cohort, including year 2020** | | | | | |
| **Up to 31 Dec. 2015** | -6.12 [-6.34, -6.04] | 1.12 [1.08, 1.14] | 0.66 [0.43, 0.76] | 2.48 [2.15, 2.62] | $\log \hat{\mathrm{D}}_x = -6.12 - 1.12 \times \log(x + 0.66) + \log(\mathrm{E}_x)$ |
| **After 31 Dec. 2015** | -5.86 [-6.09, -5.78] | 1.12 [1.08, 1.14] | 0.66 [0.43, 0.76] | 2.48 [2.15, 2.62] | $\log \hat{\mathrm{D}}_x = -5.86 - 1.12 \times \log(x + 0.66) + \log(\mathrm{E}_x)$ |
| **By cohort, excluding year 2020** | | | | | |
| **Up to 31 Dec. 2015** | -6.15 [-6.38, -6.06] | 1.12 [1.07, 1.13] | 0.65 [0.41, 0.76] | 2.22 [1.91, 2.34] | $\log \hat{\mathrm{D}}_x = -6.15 - 1.12 \times \log(x + 0.65) + \log(\mathrm{E}_x)$ |
| **After 31 Dec. 2015** | -5.88 [-6.11, -5.79] | 1.12 [1.07, 1.13] | 0.65 [0.41, 0.76] | 2.22 [1.91, 2.34] | $\log \hat{\mathrm{D}}_x = -5.88 - 1.12 \times \log(x + 0.65) + \log(\mathrm{E}_x)$ |

Supplementary Table 6: Summary table of the inferred cohort-specific model parameters with posterior median and 95% Credible intervals (Cr.I.)

##### 3.4 Forecasting

To evaluate performance on our central model, we predicted the next 200 days to assess whether the age trajectories of mortality rates were practically sensible. We assumed a stable population size over the next 200 days, and we observed substantial decreasing trends after the first year, which aligned with our assumptions (Supplementary Figures 12–15).

##### 3.5 Comparison with host country infant survival levels.

In Supplementary Table 7 we compare our infant survival estimates (right column) to those reported by the World Population Prospects (WPP) for host country populations (left column), which also include data from refugee populations (refugees are counted in their country of residence rather than country of birth). Infant survival rates for host countries are derived from the life tables estimated by WPP. We extracted data for the period 2010–2020 and calculated the average infant survival rate per country. Infant survival levels are very similar between Palestinian refugees and host country populations, particularly in Lebanon, Jordan, and Gaza. In Syria, we estimate higher survival among refugees than in the local population (98.43% vs. 98.05%). In contrast, our West Bank estimates of infant survival are higher than the State of Palestine aggregate-level figures.

| **Setting** | **Host country** (%) | **Palestine refugees** (%) |
|---|---|---|
| Jordan | 98.50 | 98.22 |
| Lebanon | 98.82 | 98.82 |
| Syria | 98.05 | 98.43 |
| State of Palestine (aggregate) | 98.41 | |
| Gaza | | 98.31 |
| West Bank | | 99.08 |

Supplementary Table 7: **Survival to age 1 among Palestine refugees and host country populations.** Infant survival rates for host countries are based on the survivor life tables estimated by World Population Prospects (WPP website). We extracted the data for the period 2010–2020 and calculated the average infant survival per country. To construct these life tables, WPP uses multiple sources including Family Health Surveys (FHS), MICS and national censuses for relevant populations and years. Data sources used in WPP life tables can be found here (WPP Data Sources).

### 4 Supplementary Figures

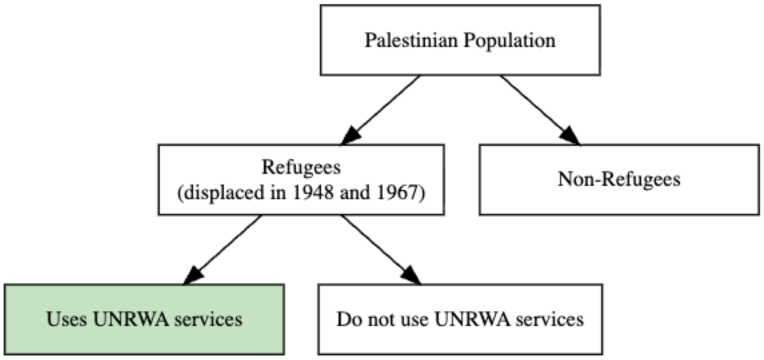

Supplementary Figure 1: The scope of UNRWA data

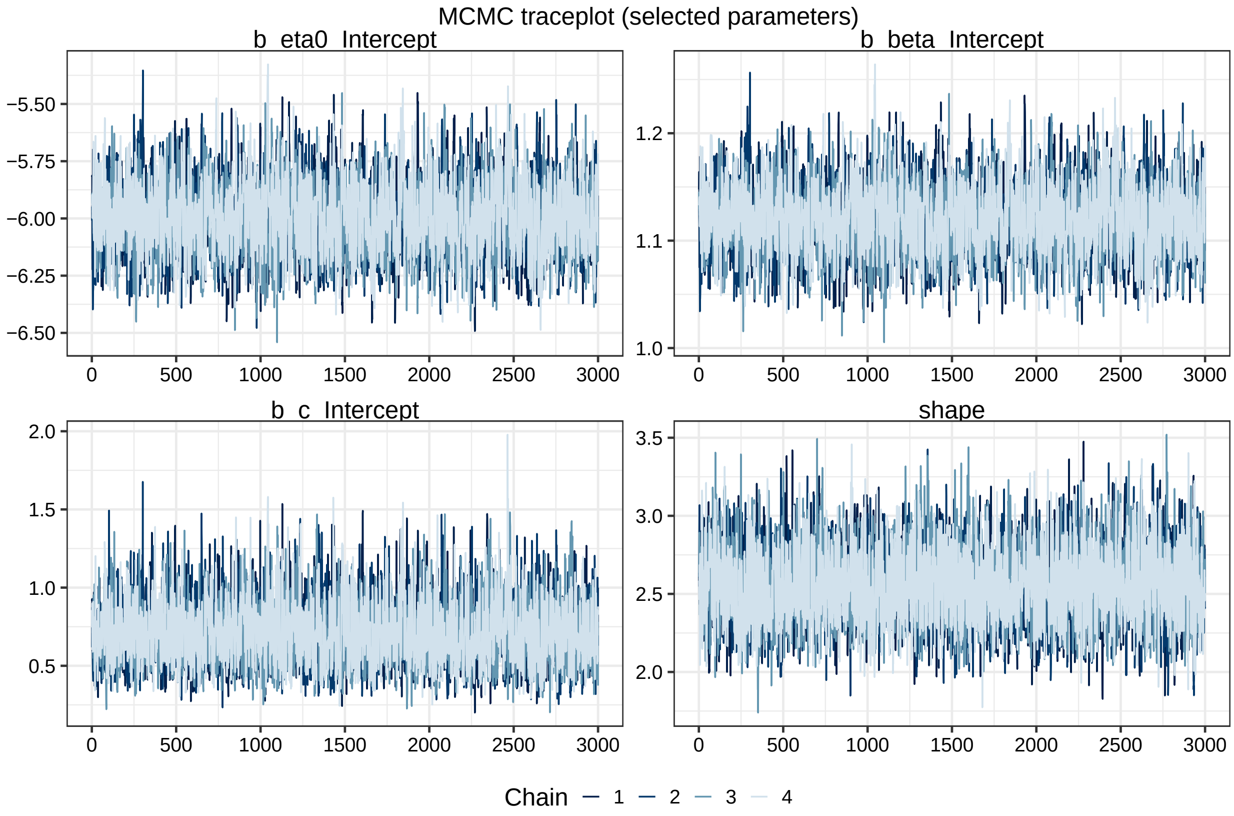

Supplementary Figure 2: **The traceplot of the model parameters of the central model.** The figure shows the trajectories of the samplers across four chains for four model parameters with the smallest number of effective sample size. Specifically, the facet names `b_eta0_Intercept`, `b_beta_Intercept`, `b_c_Intercept` and shape are $\alpha$, $\beta$, $c$ in Equation (2b)

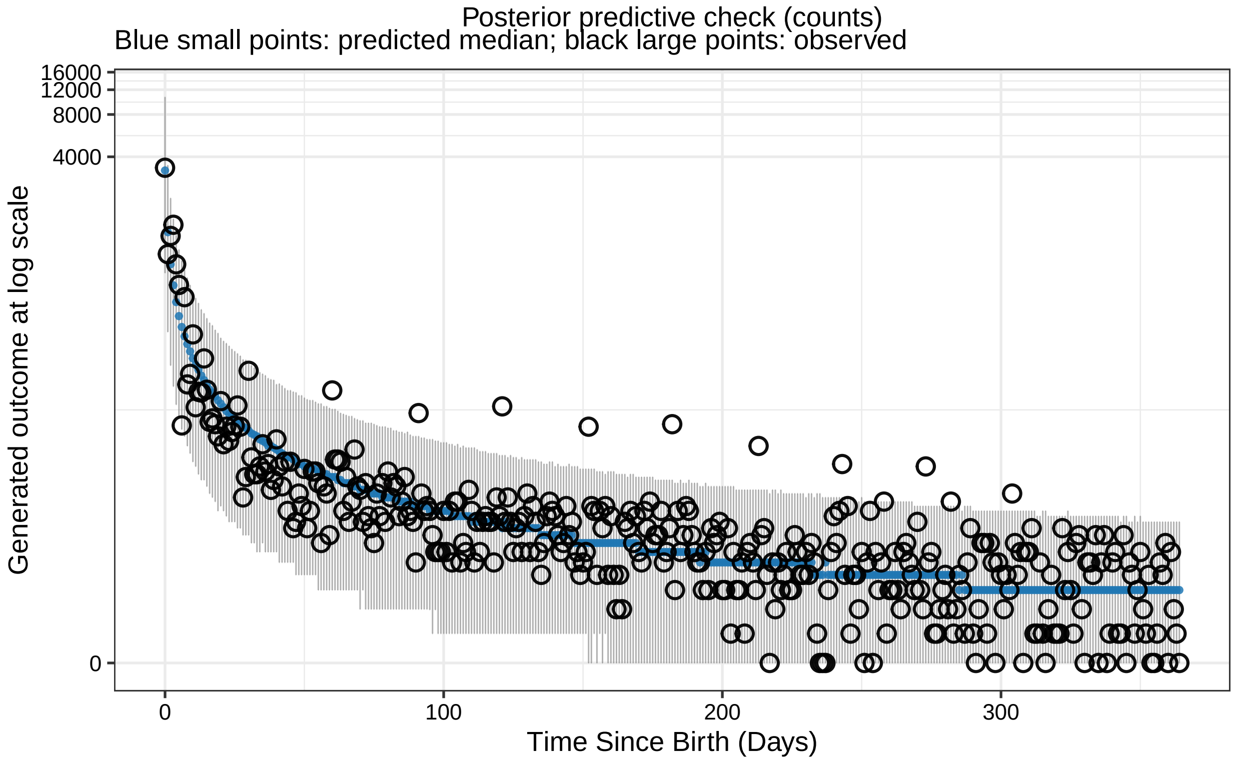

Supplementary Figure 3: **The posterior predictive checks with weights of 5.** The figure shows the observed data (black points) and the posterior median of the model predictions at log scale (blue line) with 95% P.I. (shaded area), based on the weighted likelihood of 5 in the first week.

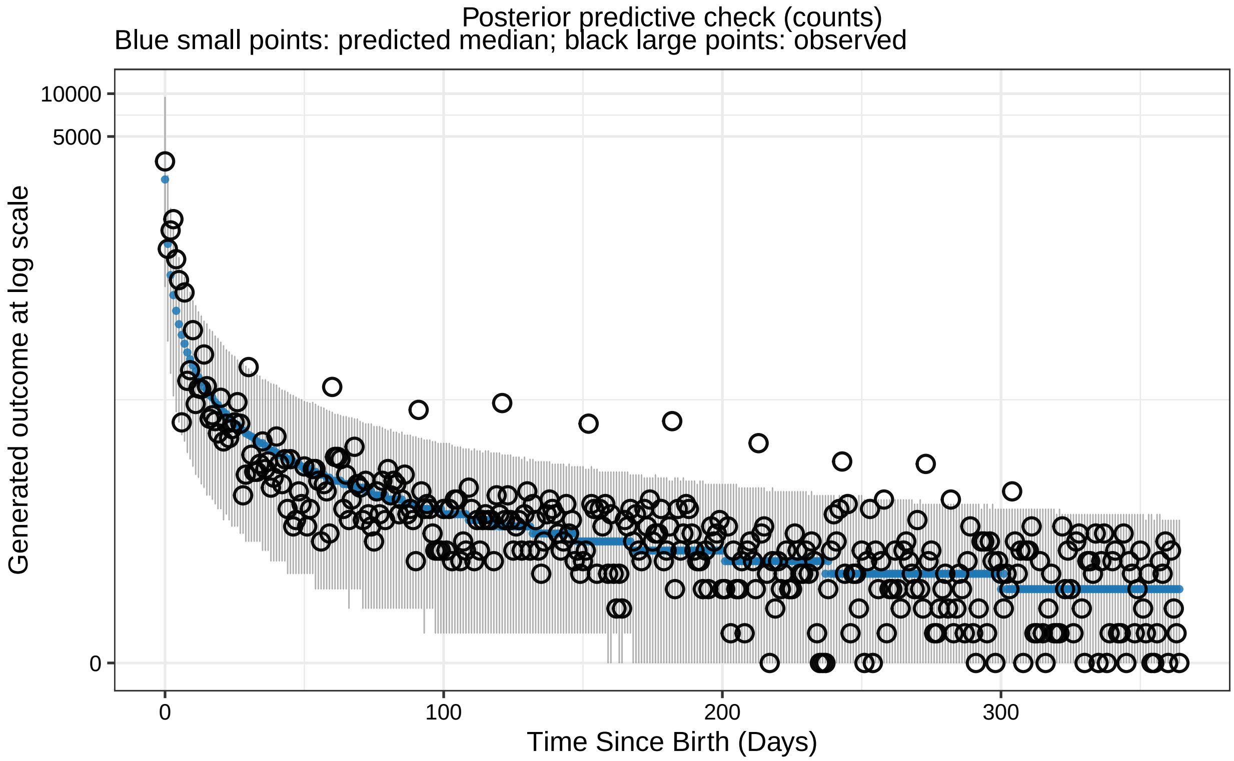

Supplementary Figure 4: **The posterior predictive checks with weights of 1.** The figure shows the observed data (black circle) and the posterior median of the model predictions at log scale (blue line) with 95% Predictive Intervals (P.I.) (shaded area), based on the weighted likelihood of 1 in the first week.

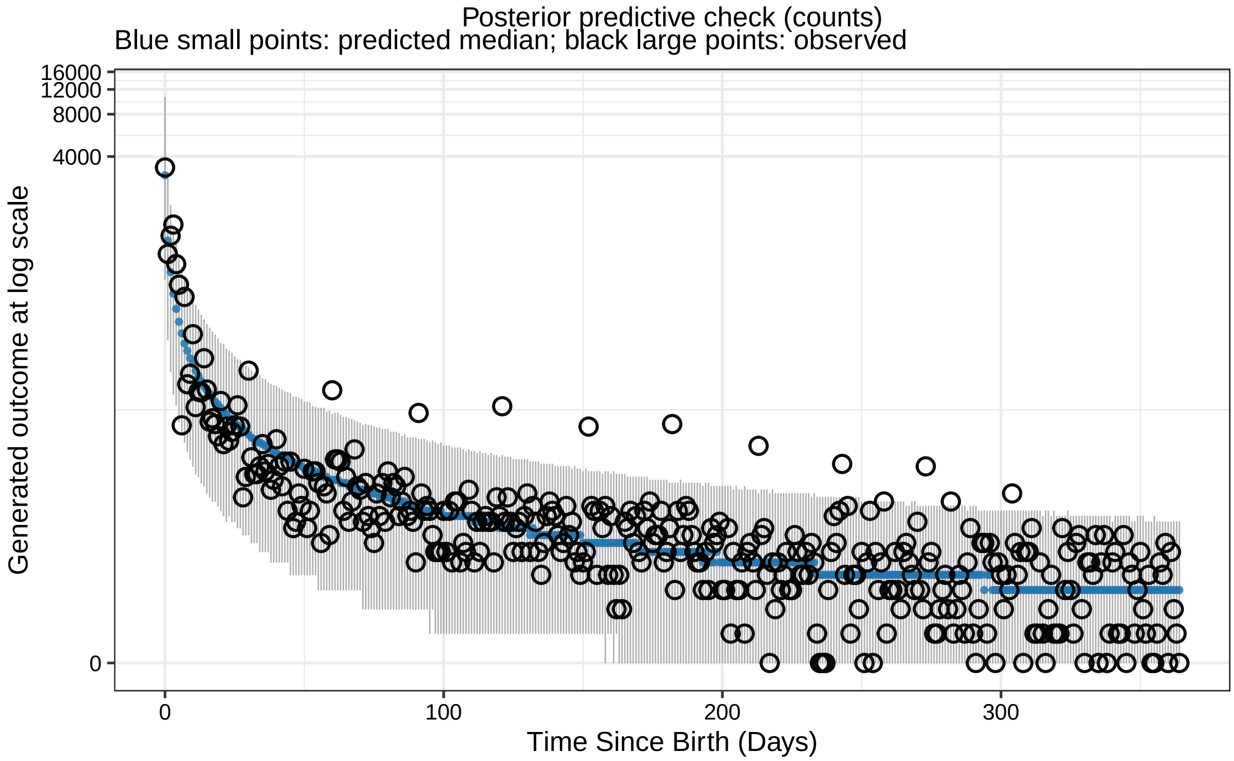

Supplementary Figure 5: **The posterior predictive checks with weights of 2.** The figure shows the observed data (black points) and the posterior median of the model predictions at log scale (blue line) with 95% P.I. (shaded area), based on the weighted likelihood of 2 in the first week.

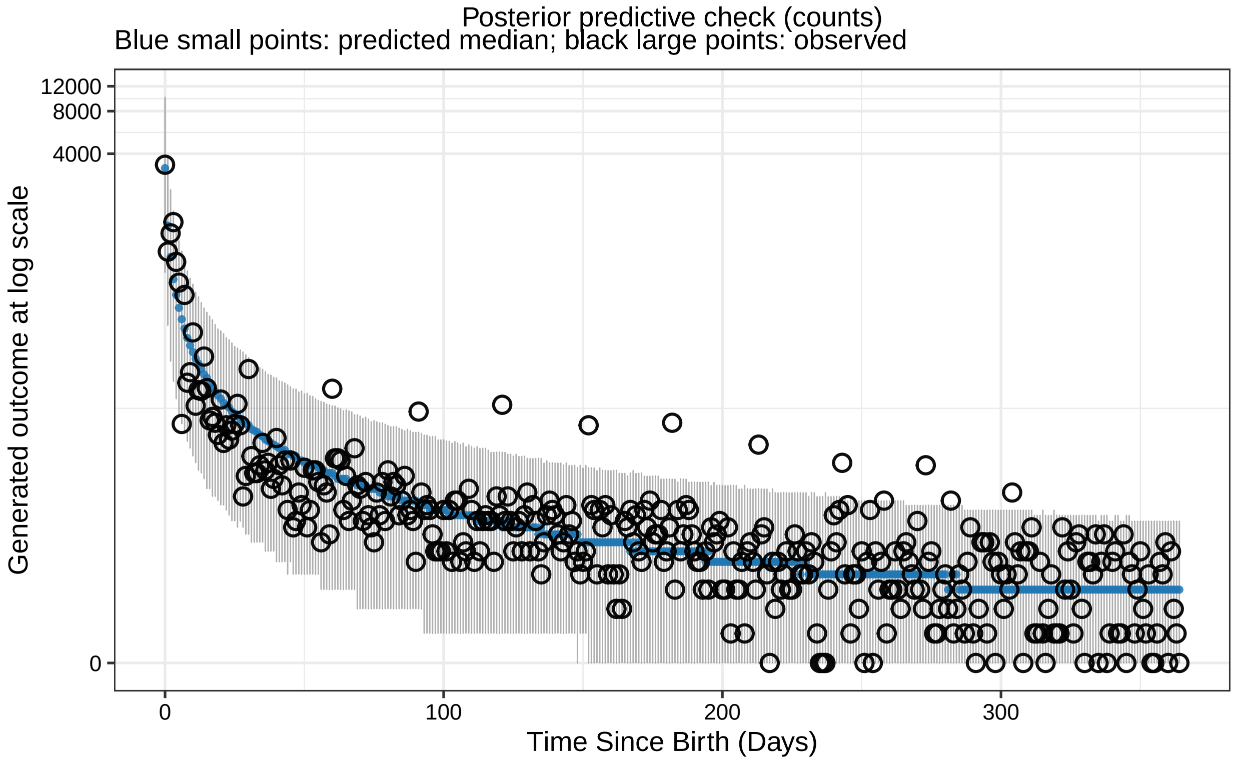

Supplementary Figure 6: **The posterior predictive checks with weights of 10.** The figure shows the observed data (black points) and the posterior median of the model predictions at log scale (blue line) with 95% P.I. (shaded area), based on the weighted likelihood of 10 in the first week.

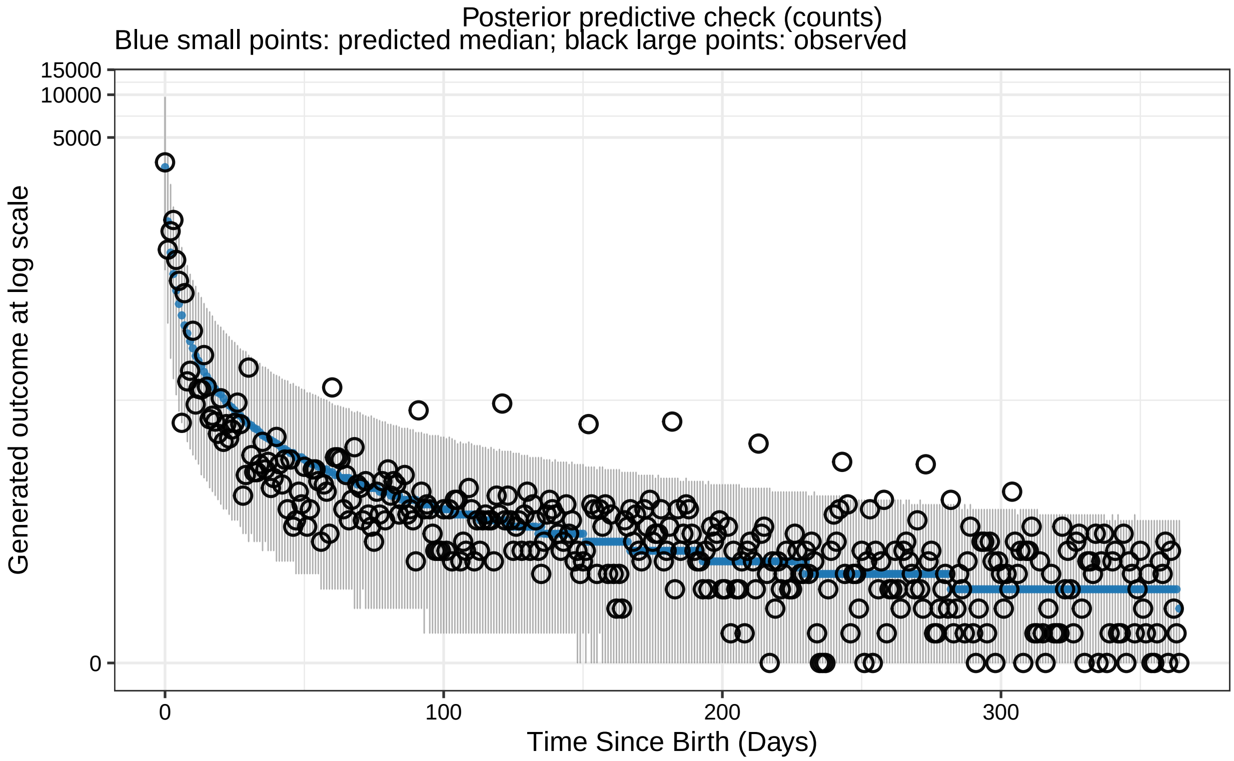

Supplementary Figure 7: **The posterior predictive checks with weights of 15.** The figure shows the observed data (black points) and the posterior median of the model predictions at log scale (blue line) with 95% P.I. (shaded area), based on the weighted likelihood of 15 in the first week.

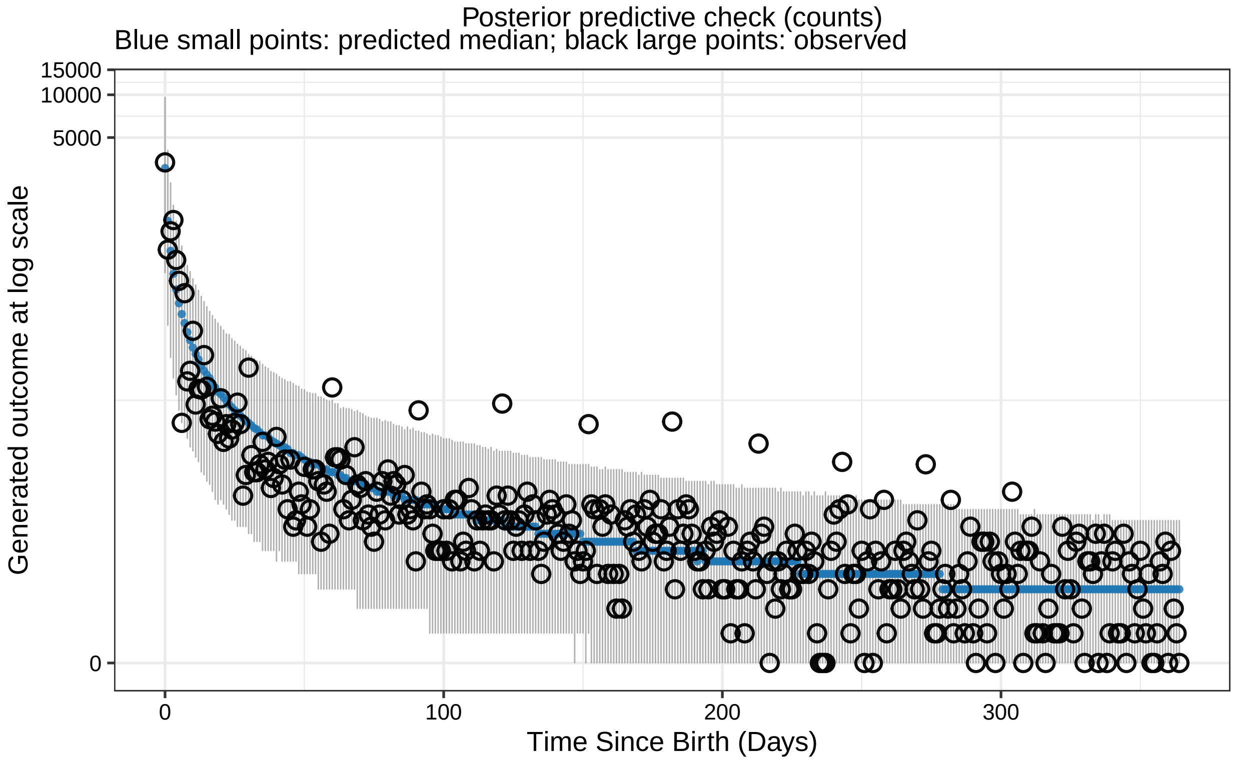

Supplementary Figure 8: **The posterior predictive checks with weights of 20.** The figure shows the observed data (black points) and the posterior median of the model predictions at log scale (blue line) with 95% P.I. (shaded area), based on the weighted likelihood of 20 in the first week.

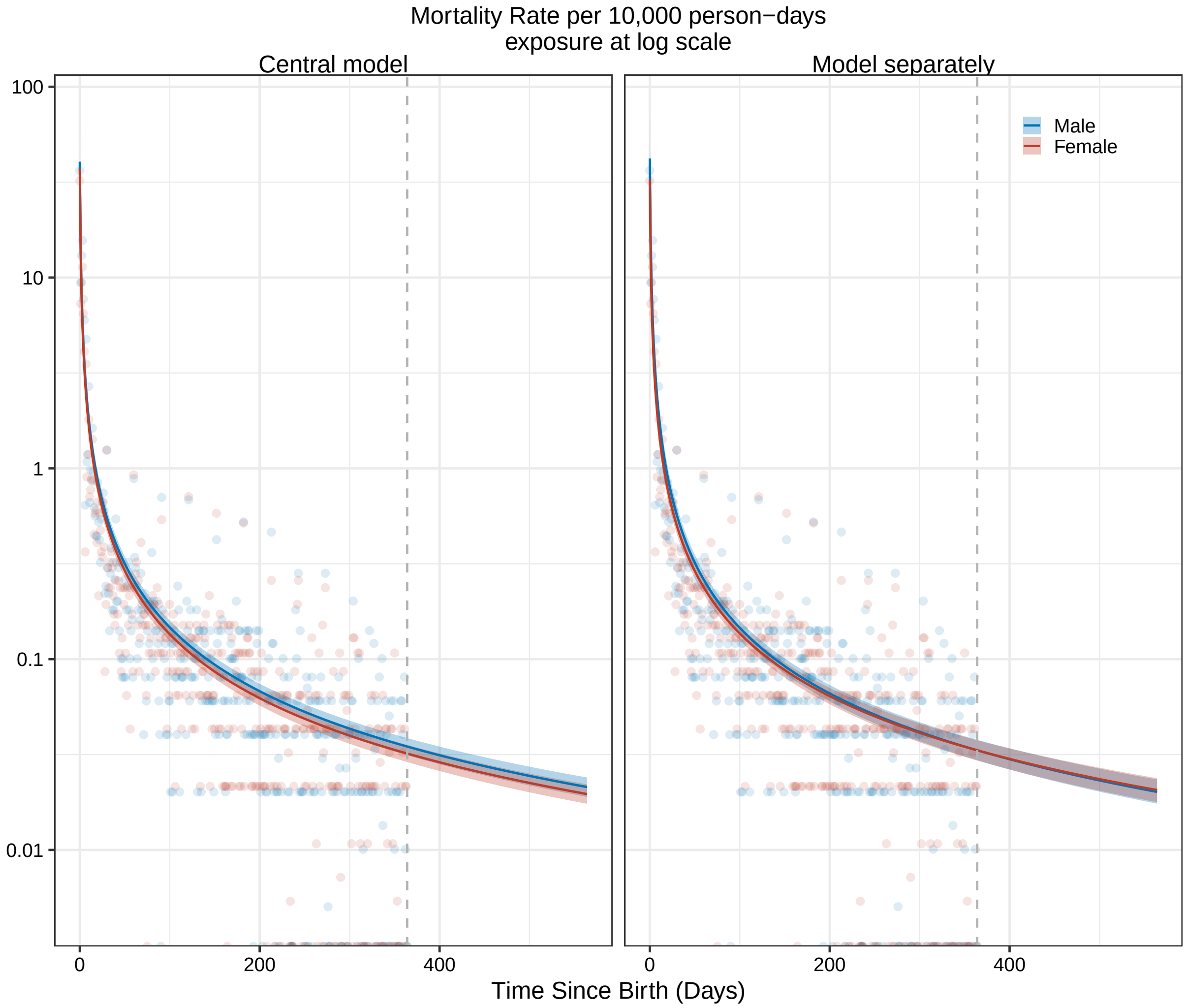

Supplementary Figure 9: **The infant mortality and survival function trends by sex in the first year since birth.** The estimated infant mortality rate per 10,000 person days with the base-10 log transformation in the first year since birth (x-axis) by sex (colour) with the fitted models with different assumptions (in column). Dashed lines represent the first year since birth.

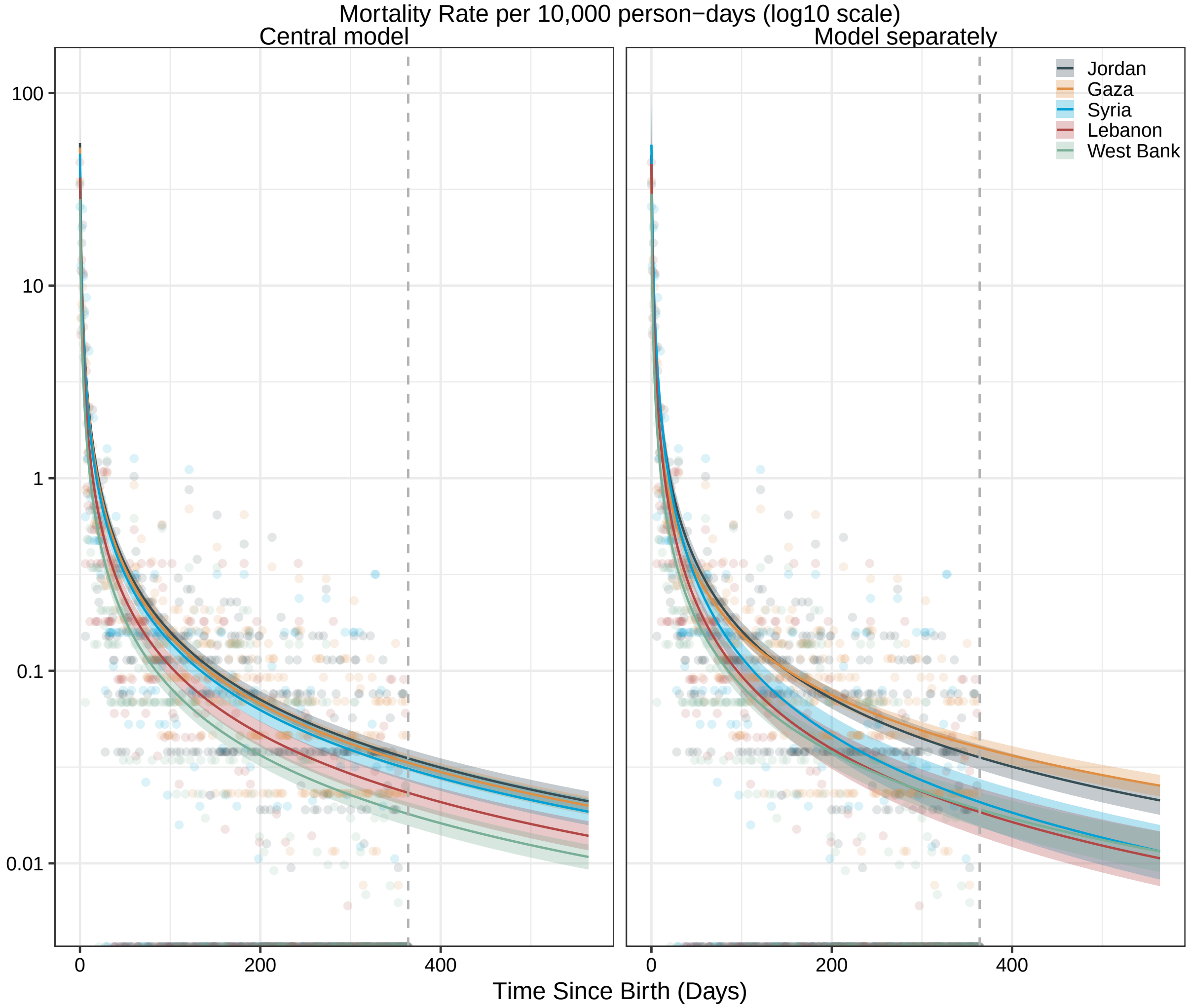

Supplementary Figure 10: **The infant mortality and survival function trends by setting in the first year since birth.** The estimated infant mortality rate per 10,000 person days with the base-10 log transformation in the first year since birth (x-axis) by setting (colour) with the fitted models with different assumptions (in column). Dashed lines represent the first year since birth.

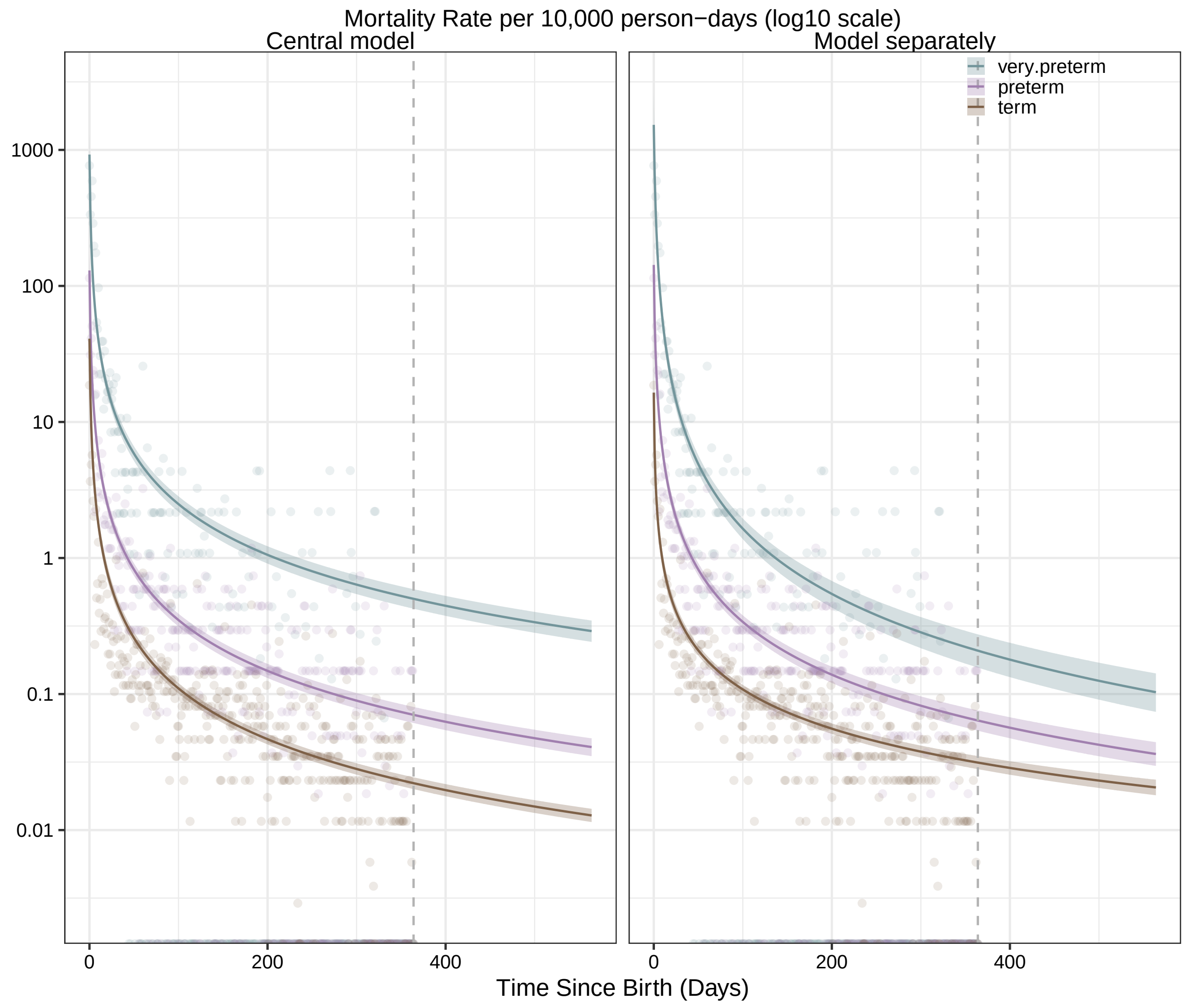

Supplementary Figure 11: **The infant mortality and survival function trends by gestational age group in the first year since birth.** The estimated infant mortality rate per 10,000 person days with the base-10 log transformation in the first year since birth (x-axis) by gestational age group (colour) with the fitted models with different assumptions (in column). Dashed lines represent the first year since birth.

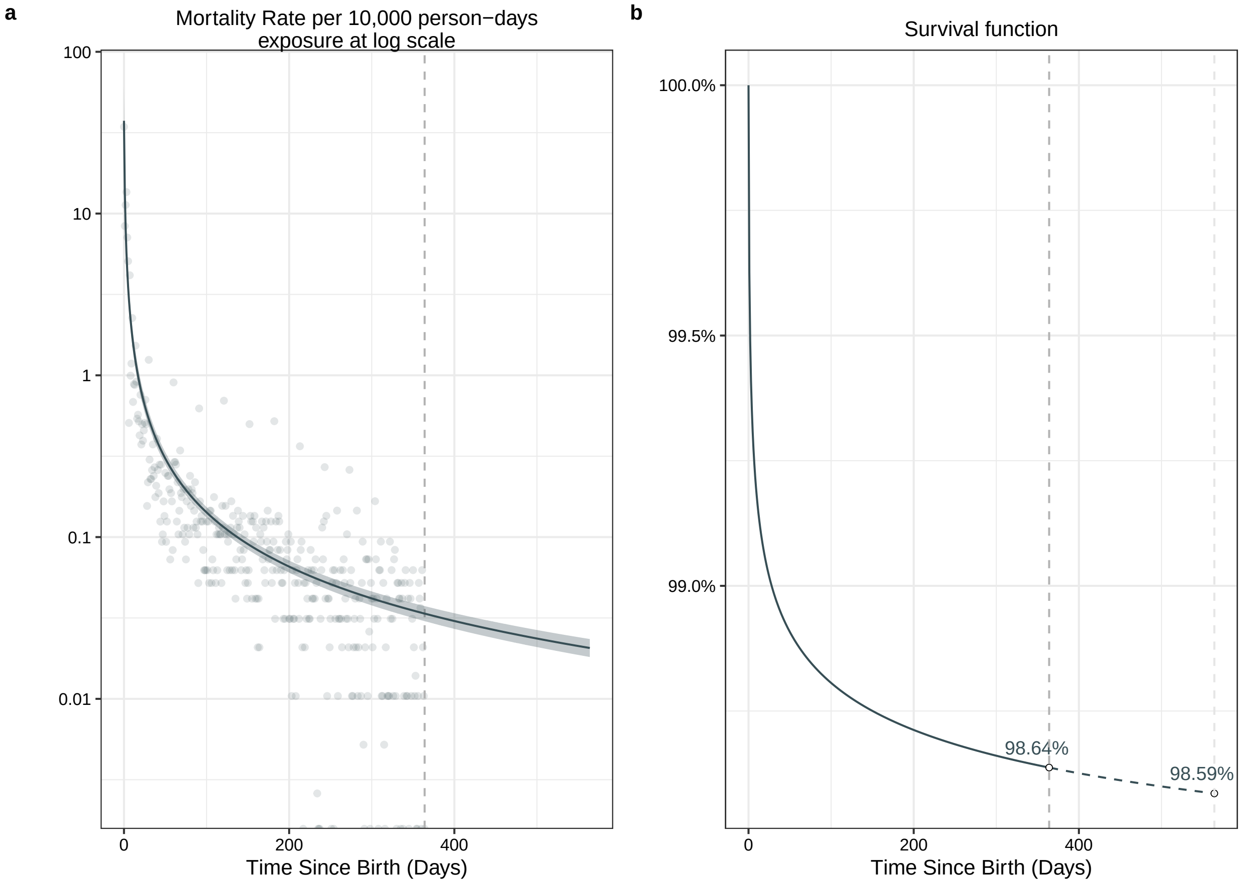

Supplementary Figure 12: **The infant mortality and survival function trends in the first 564 days since birth.** **(a).** The estimated infant mortality rate per 10,000 person days with the base-10 log transformation in the first 564 days since birth (x-axis). Dashed lines represent the first year. The posterior median values of mortality rates are shown in line with 95% Credible Interval (Cr.I.) in ribbons. The empirical mortality rates are shown in dots. **(b).** The estimated survival function based on the posterior median mortality rates. The probability of survival on day 364 and day 564 is shown in the circle with the value in text.

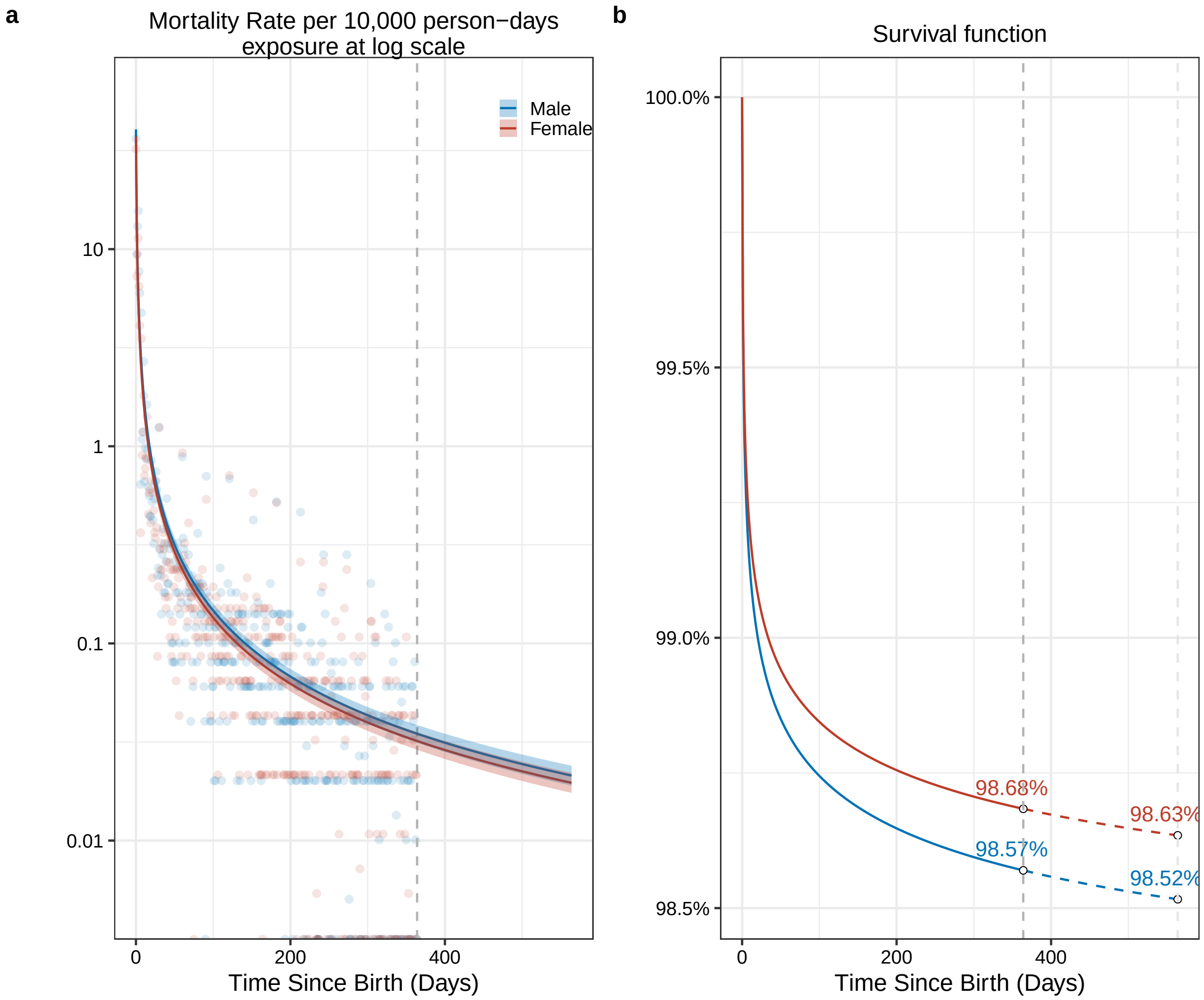

Supplementary Figure 13: **The infant mortality and survival function trends in the first 564 days by sex since birth.** **(a).** The estimated infant mortality rate per 10,000 person days with the base-10 log transformation in the first 564 days since birth (x-axis) by sex (colour). Dashed lines represent the first year. The posterior median values of mortality rates are shown in line with 95% Credible Interval (Cr.I.) in ribbons. The empirical mortality rates are shown in dots. **(b).** The estimated survival function based on the posterior median mortality rates. The probability of survival on day 364 and day 564 is shown in the circle with the value in text.

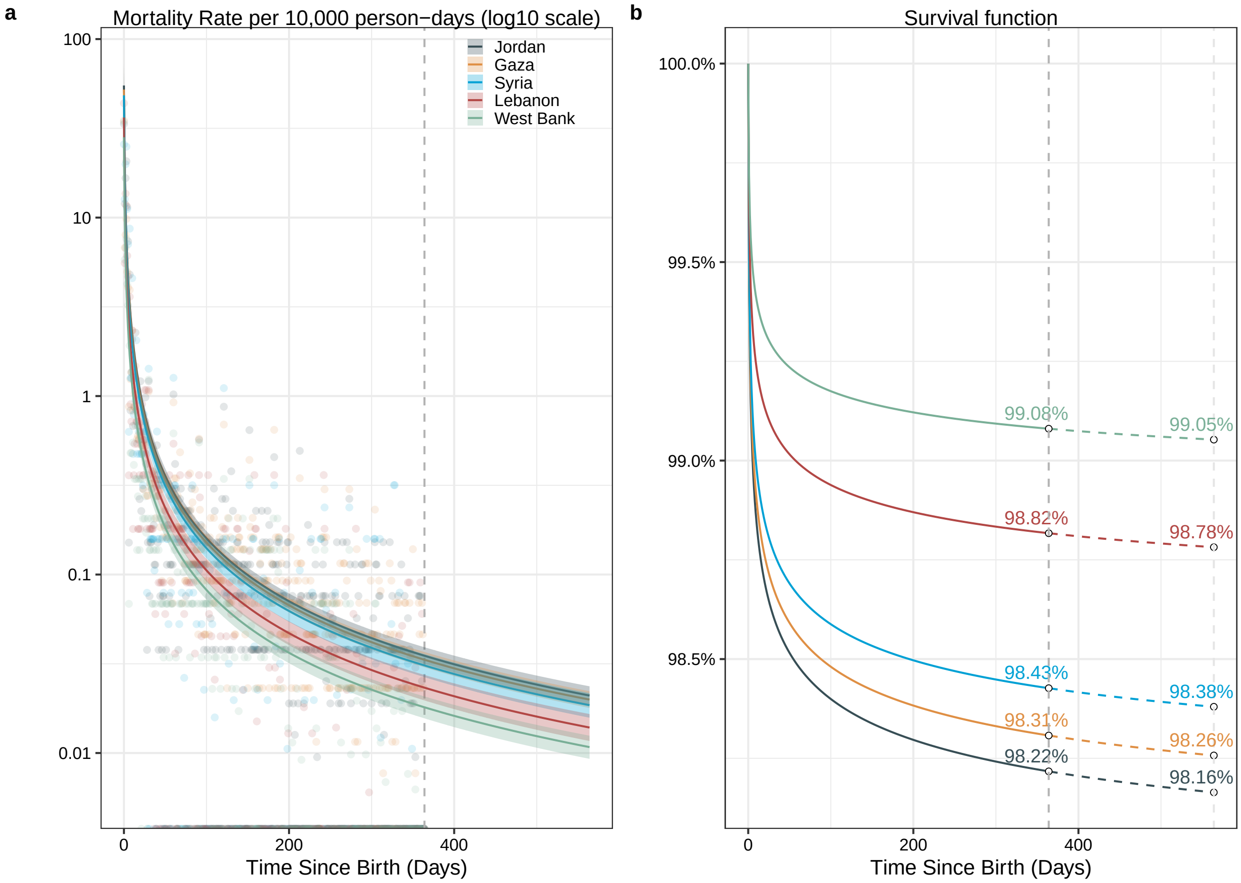

Supplementary Figure 14: **The infant mortality and survival function trends in the first 564 days by setting since birth. (a).** The estimated infant mortality rate per 10,000 person days with the base-10 log transformation in the first 564 days since birth (x-axis) by setting (colour). Dashed lines represent the first year. The posterior median values of mortality rates are shown in line with 95% Credible Interval (Cr.I.) in ribbons. The empirical mortality rates are shown in dots. **(b).** The estimated survival function based on the posterior median mortality rates. The probability of survival on day 364 and day 564 is shown in the circle with the value in text.

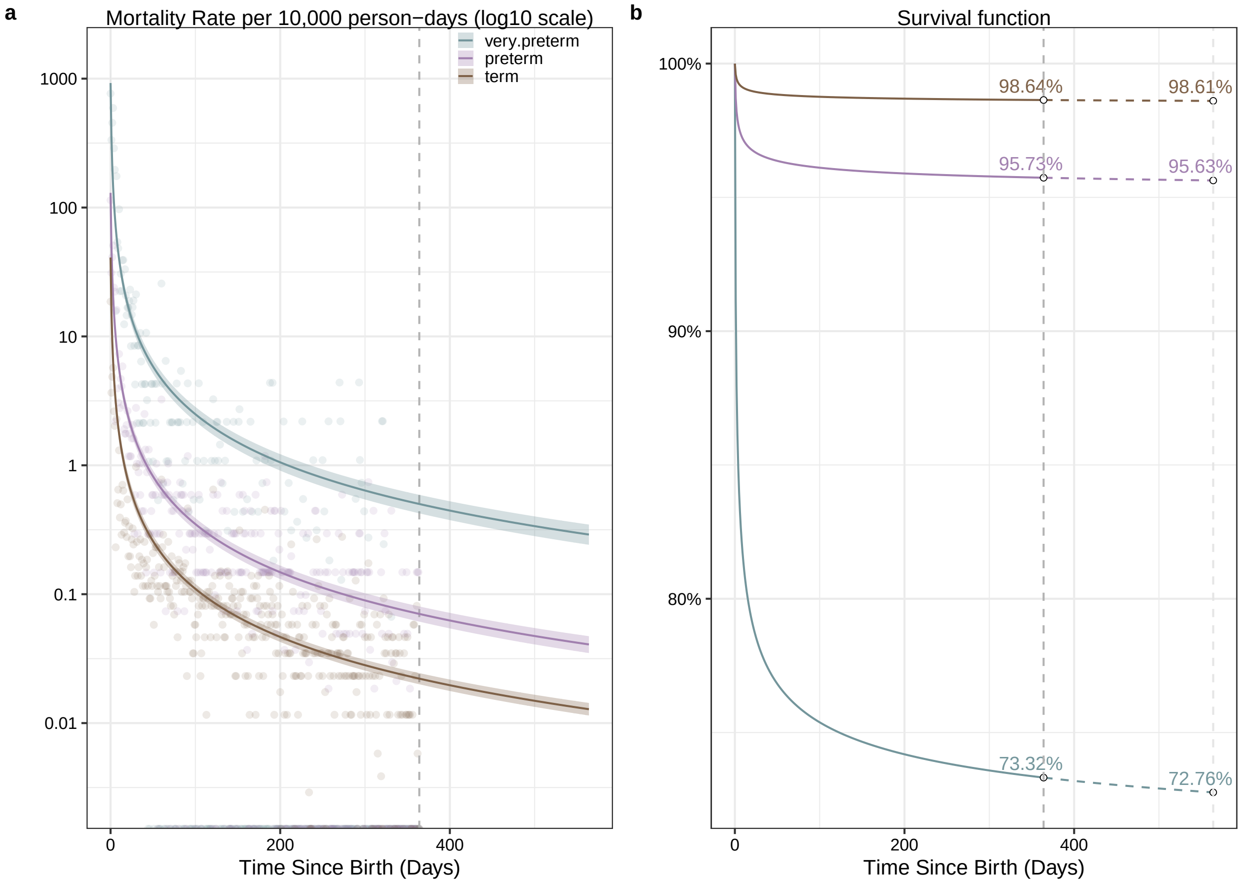

Supplementary Figure 15: **The infant mortality and survival function trends in the first 564 days by sex since birth. (a).** The estimated infant mortality rate per 10,000 person days with the base-10 log transformation in the first 564 days since birth (x-axis) by gestational age groups (colour). Dashed lines represent the first year. The posterior median values of mortality rates are shown in line with 95% Credible Interval (Cr.I.) in ribbons. The empirical mortality rates are shown in dots. **(b).** The estimated survival function based on the posterior median mortality rates. The probability of survival on day 364 and day 564 is shown in the circle with the value in text.

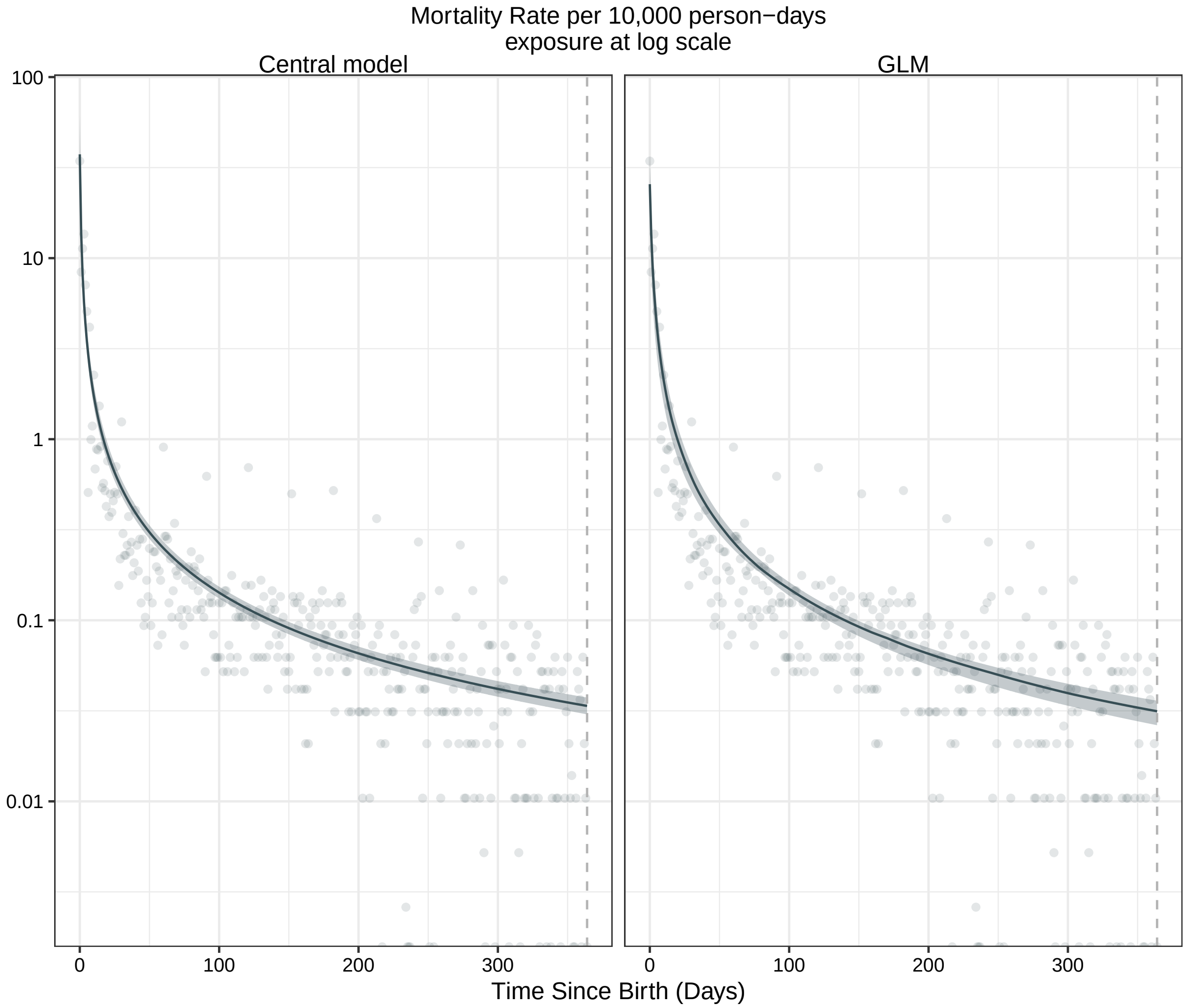

Supplementary Figure 16: **The infant mortality and survival function trends in the first year since birth, compared with results from GLM.** The estimated infant mortality rate per 10,000 person days with the base-10 log transformation in the first year since birth (x-axis) by sex (colour) with the fitted models with different model frameworks (in column). Dashed lines represent the first year since birth.

### References

Ali, M. M., Bellizzi, S. and Boerma, T. (2023), 'Measuring stillbirth and perinatal mortality rates through household surveys: a population-based analysis using an integrated approach to data quality assessment and adjustment with 157 surveys from 53 countries', *The Lancet Global Health* **11**(6), e854–e861.

Bürkner, P.-C. (2017), 'brms: An R package for Bayesian multilevel models using Stan', *Journal of statistical software* **80**, 1–28.

Khawaja, M. (2004), 'The extraordinary decline of infant and childhood mortality among palestinian refugees', *Social Science & Medicine* **58**(3), 463–470.

Riccardo, F., Khader, A. and Sabatinelli, G. (2011), 'Low infant mortality among palestine refugees despite the odds', *Bulletin of the World Health Organization* **89**(4), 304–311.

Schöley, J. (2020), The Dynamics of Ontogenescence: Modelling Age Trajectories of Feto-Infant Mortality, PhD thesis, Syddansk Universitet. `https://osf.io/cfypa/files/2ksyp`.

UNICEF State of Palestine (2021), Children in the state of palestine: Child rights brief based on findings from the 2019/2020 multiple indicator cluster survey (mics), Technical report, United Nations Children's Fund.
**URL:** *https://www.unicef.org/sop/media/1681/file/Children%20in%20the%20State%20of%20Palestine.pdf*

van den Berg, M. M., Khader, A., Hababeh, M., Zeidan, W., Pivetta, S., Abd El-Kader, M., Al-Jadba, G. and Seita, A. (2018), 'Stalled decline in infant mortality among palestine refugees in the gaza strip since 2006', *PloS one* **13**(6), e0197314.

van den Berg, M. M., Madi, H. H., Khader, A., Hababeh, M., Zeidan, W., Wesley, H., Abd El-Kader, M., Maqadma, M. and Seita, A. (2015), 'Increasing neonatal mortality among palestine refugees in the gaza strip', *PloS one* **10**(8), e0135092.
